# Specialized interferon ligand action in COVID19

**DOI:** 10.1101/2021.07.29.21261325

**Authors:** Matthew D. Galbraith, Kohl T. Kinning, Kelly D. Sullivan, Paula Araya, Keith P. Smith, Ross E. Granrath, Jessica R. Shaw, Ryan Baxter, Kimberly R. Jordan, Seth Russell, Monika Dzieciatkowska, Julie A. Reisz, Fabia Gamboni, Francesca Cendali, Tusharkanti Ghosh, Andrew A. Monte, Tellen D. Bennett, Kirk C. Hansen, Elena W.Y. Hsieh, Angelo D’Alessandro, Joaquin M. Espinosa

## Abstract

The impacts of IFN signaling on COVID19 pathology are multiple, with protective and harmful effects being documented. We report here a multi-omics investigation of IFN signaling in hospitalized COVID19 patients, defining the biosignatures associated with varying levels of 12 different IFN ligands. Previously we showed that seroconversion associates with decreased production of select IFN ligands (Galbraith et al, 2021). We show now that the antiviral transcriptional response in circulating immune cells is strongly associated with a specific subset of ligands, most prominently IFNA2 and IFNG. In contrast, proteomics signatures indicative of endothelial damage associate with levels of IFNB and IFNA6. Differential IFN ligand production is linked to distinct constellations of circulating immune cells. Lastly, IFN ligands associate differentially with activation of the kynurenine pathway, dysregulated fatty acid metabolism, and altered central carbon metabolism. Altogether, these results reveal specialized IFN ligand action in COVID19, with potential diagnostic and therapeutic implications.

**IMPACT STATEMENT:** Analysis of multi-omics signatures associated with 12 different IFN ligands reveals their specialized action in COVID19.

## INTRODUCTION

The impact of Interferon (IFN) signaling on the course of COVID19 pathology has been the subject of much investigation, with both protective and deleterious effects being reported. The protective effects of IFN signaling are demonstrated by studies showing that severe COVID19 is associated with decreased IFN signaling (1), the presence of auto-antibodies blocking IFN ligand action (2), and genetic variants that impair IFN signaling (3). However, Type I IFN signaling has been established as a driver of pathology in mouse models of both SARS-CoV-1 and SARS-CoV-2 infections (4, 5), and Type I and III IFN signaling have been implicated in disruption of lung barrier function and increased susceptibility to secondary bacterial infections in mice (6). This ambivalence has fueled the design of seemingly contradictory clinical trials using either IFN ligands (7) or agents that block IFN signaling, such as JAK inhibitors (8). This duality is further illustrated by studies showing that genetic variants leading to low expression of the Type I IFN receptor IFNAR2 or high expression of TYK2, a protein kinase required for Type I IFN signaling, are associated with life-threatening disease (9). Therefore, it is possible that context-dependent variations in IFN signaling may attenuate or exacerbate COVID19 pathology in different settings. Indeed, retrospective analysis of IFN-α2b treatment in COVID19 showed that early administration was associated with reduced mortality, whereas late administration was associated with increased mortality (10).

There are three major types of IFN signaling defined by the transmembrane receptors and downstream signaling kinases involved (11). Type I IFN involves IFN alpha, beta, epsilon, kappa, and omega ligands, the IFNAR1 and IFNAR2 receptors, and the downstream kinases JAK1 and TYK2. Type II IFN signaling involves the gamma ligand, the IFNGR1 and IFNGR2 receptors, and the downstream kinases JAK1 and JAK2. Type III IFN signaling involves the lambda ligands, the IFNLR1 and IL10RB receptors, and the JAK1 and TYK2 kinases. However, this classification fails to capture the biological complexity created by the differential effects of distinct ligands acting through the same receptors. This is most evident by the differential effects of alpha ligands and IFNB1 within Type I signaling (12). Even within alpha ligands there is significant heterogeneity in cellular source, site of action, and downstream effects (12). Nevertheless, in the context of lung viral infections, it is accepted that select alpha and lambda ligands are first responders in the antiviral response due to their induced expression upon engagement of pattern recognition receptors in the lung epithelium (13). In the context of SARS-CoV-2 infections, little is known about the functional specialization of different IFN ligands and their relative contributions to different aspects of the ensuing pathology. Furthermore, SARS-CoV-2 has evolved diverse strategies to evade IFN signaling (14), and clinical trials for IFN alpha, beta, gamma and lambda ligands have been completed or are under way, even in combinations, but definitive results leading to approval for clinical use are pending (15).

Within this context, we report here a multi-omics analysis of IFN signaling in hospitalized COVID19 patients. This investigation includes a comprehensive examination of the whole blood transcriptome, plasma proteome, anti-SARS-CoV-2 antibodies, peripheral immune cell repertoire, and plasma and red blood cell metabolomes in relationship to levels of 12 different circulating IFN ligands. In hospitalized patients with moderate COVID19 pathology, transcriptome-based IFN scores are highly variable and significantly associated with levels of a subset of circulating IFN ligands such as IFNA2 and IFNG, but not so IFNA6 or IFNB1. Likewise, plasma proteomic signatures are also differential among ligands. For example, whereas IFNG and other ligands are clearly associated with production of monocyte activating and mobilizing chemokines, IFNA6 and IFNB1 levels associate with markers of platelet degranulation and endothelial damage. Furthermore, IFN ligands display differential relationships with immunoglobulins targeting SARS-CoV-2, revealing that seroconversion associates with decreased production of a select subset of ligands. This shift in IFN ligand production upon seroconversion is accompanied by significant changes in the immune cell types associated with production of the various ligands. For example, whereas IFNA10 is strongly associated with levels of Th1 CD4 T cells, CD56^bright^ NK cells and plasmacytoid dendritic cells, its levels are strongly anti-correlated with levels of circulating plasmablasts. Lastly, we revealed specific metabolomic signatures associated with diverse ligands.

Whereas IFNG is the most strongly associated with tryptophan catabolism through the kynurenine pathway, other ligands associate with metabolic pathways indicative of dysregulated central carbon metabolism, nitric oxide metabolism, and fatty acid oxidation. Altogether, these results indicate that modulation of IFN signaling in the clinic, either with agonists or antagonists, must take into account the endogenous state of the IFN ligand milieu, and that ligand-specific effects are to be expected.

## RESULTS

### Variable IFN signaling in COVID19 associates with levels of a specific subset of IFN ligands

In this study, we analyzed the multi-omics datasets generated by the COVIDome Project (covidome.org) to investigate IFN signaling in hospitalized COVID19 patients. The COVIDome Project datasets have been previously described (16, 17) and include matched analysis of whole blood transcriptome, plasma proteomics via complementary SOMAscan^®^ assays and mass spectrometry, measurement of 82 immune factors by multiplexed immunoassays, SARS-CoV-2 seroconversion assays, immune cell profiling by mass cytometry, plasma and red blood cell metabolomics, as well as annotated clinical data. The COVIDome Project cohort analyzed in this study consists of 73 hospitalized COVID19 patients with mild-to-moderate disease versus 32 controls (see **Supplementary file 1** for cohort characteristics and **Methods**). To monitor IFN signaling in this cohort, we first analyzed the transcriptome dataset. Using DESeq2 analysis, with adjustment for age and sex as covariates, we identified 2299 genes differentially expressed in the blood of COVID19 patients (**Figure 1A, Supplementary file 2**). Gene Set Enrichment Analysis (GSEA) identified the Hallmark Interferon Alpha and Gamma Response gene sets as the most significant positively enriched gene signatures in COVID19 patients (**Figure 1B, Supplementary file 3**). To assess inter-individual variation in expression of these IFN gene signatures, we calculated Z-score-based ‘IFN Alpha’ and ‘IFN Gamma’ transcriptional scores for each sample, showing that COVID19 patients show significantly increased yet highly variable IFN scores relative to controls (**Figure 1C**, **Figure 1 – supplement 1A**). In order to assess the degree to which this variability in IFN signaling was associated with the levels of circulating IFN ligands, we mined the SOMAscan^®^ proteomics and multiplexed immunoassay datasets (Meso Scale Discovery assays, MSD, see **Methods**), which collectively measured a total of 17 different IFN ligands. To test the specificity of the reagents measured in these two platforms, we spiked single concentrations (SOMAscan^®^) or multiple concentrations (MSD) of 16 of these IFN ligands, for which purified recombinant proteins were commercially available, into a pooled plasma reference sample (only IFNW1 could not be obtained, see **Methods**). This test led us to discard five SOMAscan^®^ measurements (IFNA5, IFNA8, IFNA14, IFNA21, IFNL2) due to lack of sensitivity, and to relabel some measurements based on apparent cross-reactivity, such as IFNA4/16, IFN7/17/21, and IFNL3/2 (**Figure 1 – supplement 1B**). When the same IFN ligand was measured by both platforms, we preferred the MSD measurements, which are quantified against a standard curve (**Figure 1 – supplement 1C**). This exercise allowed us to focus on measurements for 12 IFN ligands in our subsequent analyses: IFNA1, IFNA2, IFNA4/16, IFNA6, IFNA7/17/21, IFNA10, IFNA16, IFNB1, IFNG, IFNL1, INFL3/2, and IFNW1 (**Figure 1 – supplement 1B-D**). We next determined Spearman correlations between the RNA-based IFN Alpha transcriptional scores and levels of the 12 IFN ligands (**Figure 1D**). Interestingly, the correlations were highly variable, and four of the ligands did not show significant associations with the IFN Alpha transcriptional scores (IFNB1, IFNA16, IFNW1, and IFNA6). This result is clearly illustrated by the Type I ligands IFNA2 and IFNA6, which are the most and least correlated with IFN Alpha scores, respectively. Although both ligands are significantly upregulated in the plasma of COVID19 patients (**Figure 1E**), only IFNA2 levels correlate with the IFN Alpha scores (**Figure 1F**) and with expression of well recognized ISGs, such as ISG15 and OAS2 (**Figure 1F-G**).

**Figure 1.**
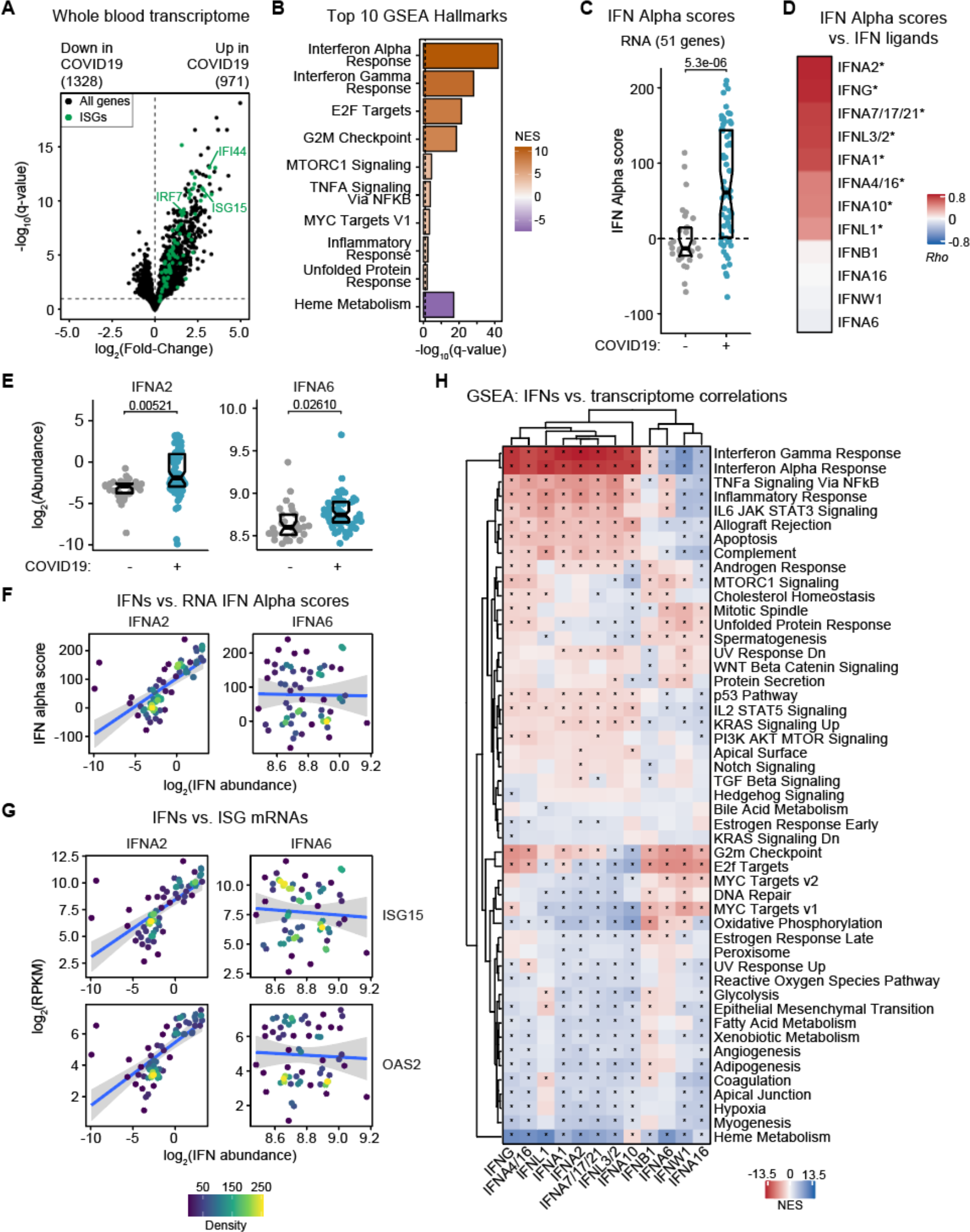
IFN signaling at the whole-blood transcriptome level correlates with a subset of IFN ligands. (**A**) Volcano plot for DESeq2 differential expression analysis of gene-level count data for COVID19-positive vs. -negative samples, adjusted for age and sex. Horizontal dashed line indicates an FDR threshold of 10% for negative binomial Wald test; numbers above plot indicate significant genes at this threshold. Interferon stimulated genes (ISGs) are highlighted in green. (**B**) Bar plot of top 10 Hallmark gene sets as ranked by absolute normalized enrichment score (NES) from Gene Set Enrichment Analysis (GSEA). Bar color represents NES; Bar length represents -log10(FDR q-value). (**C**) RNA-based IFN Alpha scores, separated by COVID19 status. Scores were calculated for each research participant by summing Z-scores for 51 differentially expressed genes from the Interferon Alpha Response Hallmark gene set from MSigDB. Z-scores were calculated from the adjusted concentration values for each gene in each sample, based on the mean and standard deviation of COVID19-negative samples. Data are presented as a modified sina plot with box indicating median and interquartile range. (**D**) Ranked heatmap representing correlations between RNA-based IFN Alpha scores and plasma levels of each IFN ligand. Values displayed are Spearman correlation coefficients (*Rho*); asterisks indicate significant correlations (10% FDR). (**E**) Sina plots comparing abundance for the indicated IFNs in COVID19-negative (-) vs. -positive (+) plasma samples. Data are presented as modified sina plots with boxes indicating median and interquartile range. Numbers above brackets are q-values for Mann–Whitney tests. (**F**) Scatter plots showing the relationship between RNA-based IFN Alpha score and plasma abundance of the indicated IFNs in COVID19-positive patients. Points are colored by density; blue lines represent linear model fit with 95% confidence intervals in grey. (**G**) Scatter plots showing the relationship between ISG mRNA levels and plasma abundance of the indicated IFNs in COVID19-positive patients. Points are colored by density; blue lines represent linear model fit with 95% confidence intervals in grey. (**H**) Heatmap representing enrichment of Hallmark gene sets among Spearman correlations between mRNA levels and plasma levels of each IFN ligand. Values displayed are NES from GSEA; asterisks indicate significant enrichment (10% FDR); columns and rows are grouped by hierarchical clustering. See also **Figure 1 – supplement 1** and **Figure 1 – supplement 2**.

These differences could not be simply explained by the degree of induction of the various ligands in COVID19 patients, as illustrated by IFNA7/17/21, IFNL3/2, IFNA10 and IFNL1, all of which were not statistically higher among COVID19 patients (**Figure 1 – supplement 1D**) but nonetheless correlated significantly with IFN Alpha scores (**Figure 1D**). Repeating this analysis for IFN Gamma scores produced a very similar rank of correlations, which is perhaps not surprising given the high overlap between the IFN Alpha and Gamma Response Hallmark GSEA gene sets (18) (**Figure 1 – supplement 1A**, see **Methods**).

To explore this phenomenon more deeply, we completed a comprehensive analysis of gene expression signatures in the whole blood transcriptome associated with varying plasma levels of the 12 IFN ligands, using only data from COVID19 patients. Toward this end, we defined Spearman correlations between each ligand and mRNAs for 15,000+ genes detected by RNAseq, which identified thousands of significant positive and negative correlations, with great variability across ligands (**Figure 1 – supplement 2A, Supplementary file 4**). We then analyzed the ranked correlations for each IFN ligand using GSEA to identify known gene sets with significant enrichment among positive or negative correlations with the levels of each ligand (**Figure 1H, Supplementary file 5**). This analysis showed that the top gene signatures positively associated with 8 of the IFN ligands are indeed the IFN Alpha and Gamma Responses, followed by related inflammatory and immune pathways (e.g. TNFA signaling, Inflammatory response, IL6/JAK/STAT3 signaling). In contrast, for the other 4 ligands (IFNB1, IFNA6, IFNW1, and IFNA16), the top signatures enriched in the positive correlations are related to cell proliferation, such as G2M checkpoint, E2F targets, and MYC targets (**Figure 1H**). In fact, some of these ligands show negative correlations with the IFN Alpha and Gamma Responses (**Figure 1H**). This result is once again illustrated by the differential behavior of IFNA2 and IFNA6. Whereas mRNAs positively associated with IFNA2 show clear enrichment of the IFN Alpha Response gene set, these same mRNAs are negatively correlated with IFNA6 levels (e.g. ISG15, **Figure 1 – supplement 2B**). Altogether, these results suggest functional specialization among circulating IFN ligands, whereby only a fraction of ligands associates with the recognizable IFN transcriptional response in circulating immune cells.

**Figure 2.**
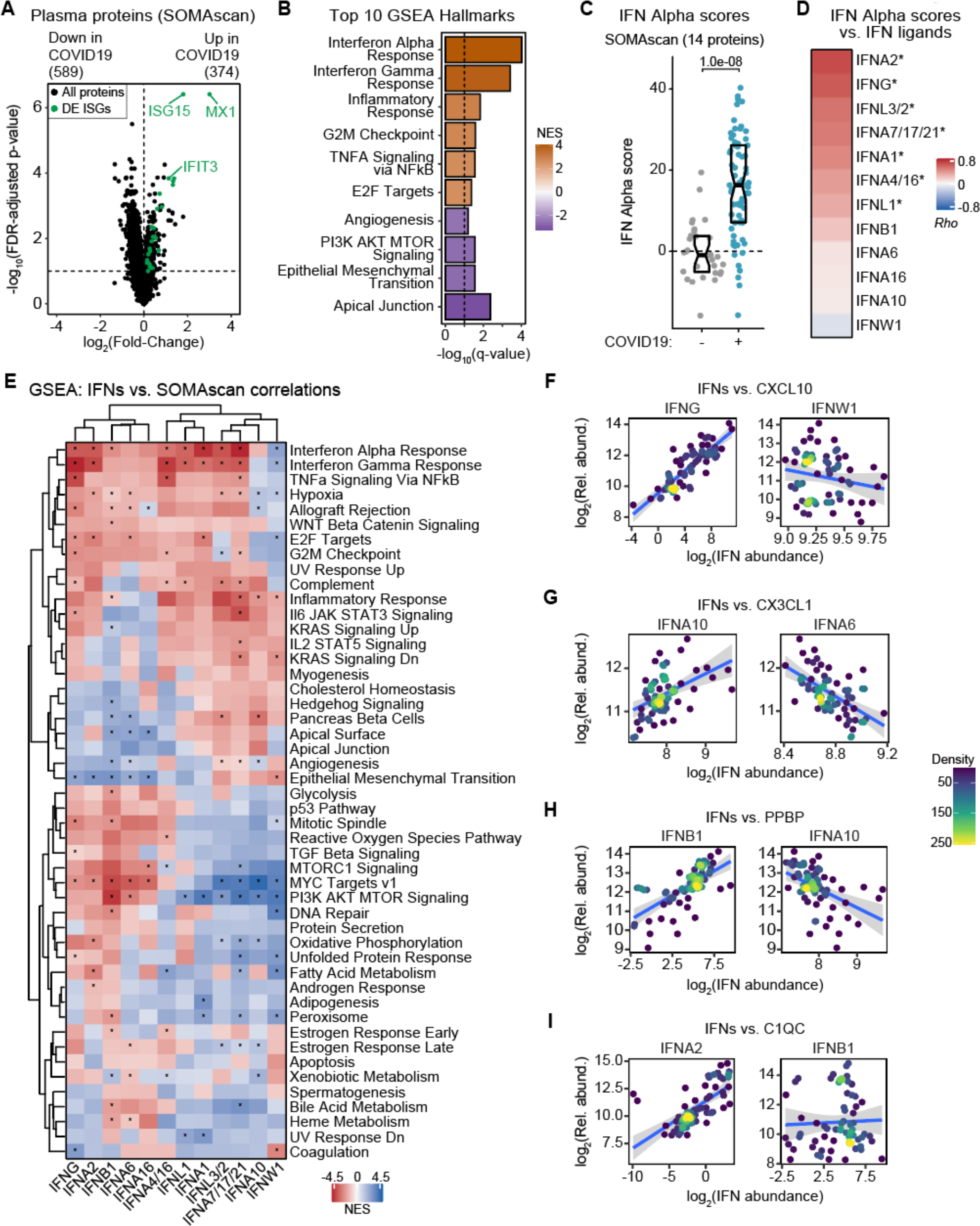
IFN signaling at the proteome level correlates with features of COVID19 pathophysiology. (**A**) Volcano plot for linear regression analysis of Somascan plasma protein abundance data for COVID19-positive vs. -negative samples, adjusted for age and sex. Horizontal dashed line indicates an FDR threshold of 10% (q < 0.1); numbers above plot indicate significant genes at this threshold. Proteins encoded by Interferon Stimulated Genes (ISGs) are highlighted in green. (**B**) Barplot of top 10 Hallmark gene sets as ranked by absolute normalized enrichment score (NES) from Gene Set Enrichment Analysis (GSEA). Bar color represents NES; Bar length represents -log10(FDR q-value). (**C**) Protein-based IFN Alpha scores, separated by COVID19 status. Scores were calculated for each research participant by summing Z-scores for 14 differentially abundant proteins from the Interferon Alpha Response Hallmark gene set from MSigDB. Z-scores were calculated from the adjusted concentration values for each gene in each sample, based on the mean and standard deviation of COVID19-negative samples. Data are presented as a modified sina plot with box indicating median and interquartile range. (**D**) Ranked heatmap representing correlations between protein-based IFN Alpha scores and plasma levels of each IFN ligand. Values displayed are Spearman correlation coefficients (*Rho*); asterisks indicate significant correlations (10% FDR). (**E**) Heatmap representing enrichment of Hallmark gene sets among Spearman correlations between plasma levels of proteins measured by SOMAscan and each IFN ligand. Only proteins with at least one significant correlation are shown. Values displayed are NES from GSEA; asterisks indicate significant enrichment (10% FDR); columns and rows are grouped by hierarchical clustering. (**F-I**) Scatter plots comparing relationships between plasma proteins and the indicated IFNs in COVID19-positive patients. Points are colored by density; blue lines represent linear model fit with 95% confidence intervals in grey. See also Figure 2 **– supplement 1** and Figure 2 **– supplement 2**.

### IFN ligands show differential proteomic signatures associated to COVID19 pathophysiology

Next, we investigated the proteomic signatures associated with each ligand. Using a linear model adjusting for age and sex, we identified 963 epitopes measured by the SOMAscan^®^ platform differentially abundant in the plasma of COVID19 patients (**Figure 2A, Supplementary file 6**). GSEA identified Hallmark IFN Alpha and Gamma Responses as the top proteomic signatures induced in COVID19 (**Figure 2B, Supplementary file 7**). As for the transcriptome, we created protein-based IFN alpha and gamma scores for each participant, which showed significantly higher yet highly variables IFN scores among COVID19 patients (**Figure 2C**, **Figure 2 – supplement 1A**). Notably, plasma protein-based IFN scores may inform about the organismal IFN response, not just that of circulating immune cells driving the whole blood transcriptome IFN signature, as multiple organs and tissues could contribute to secretion of IFN-related proteins. We then defined correlations between the 12 ligands and the protein-based IFN scores, which revealed some similarities and differences relative to the RNA-based IFN scores (**Figure 2D**). Whereas IFNA2 and IFNG remained as the ligands most correlated with the protein-based IFN Alpha and Gamma scores, other ligands behaved differently (**Figure 2D**, **Figure 2 – supplement 1A**). For example, IFNA10, which was significantly correlated with the RNA-based IFN Alpha and Gamma scores, was not so with the protein-based IFN Alpha and Gamma scores. In contrast, IFNA6 and IFNB1 ranked higher in their association with the protein-based scores (**Figure 1D**, **Figure 2D**, **Figure 2 – supplement 1A**). As for the transcriptome analysis, we then defined Spearman correlations between each of the 12 ligands and 4800+ epitopes measured by SOMAscan^®^ proteomics and analyzed the correlation results with GSEA (**Figure 2 – supplement 1B**, **Figure 2E**, **Supplementary file 8, Supplementary file 9**). Interestingly, some IFN ligands with weak transcriptome signatures nonetheless present strong proteomic signatures. For example, IFNA6 and IFNB1, which show very weak correlations with mRNAs (**Figure 1 – supplement 2A**), are among the ligands with the most numerous significant associations with circulating proteins (**Figure 2 – supplement 1B**). In fact, unsupervised hierarchical clustering of the proteomic GSEA signatures placed IFNA6 and IFNB1 together with IFNA2 and IFNG (**Figure 2E**). This suggest that whereas IFNA6 and IFNB1 may not contribute to the IFN transcriptional response of circulating immune cells, they may nonetheless contribute to IFN responses in peripheral tissues and organs contributing to the protein-based plasma IFN signature. This is illustrated by the behavior of CXCL11 (IFN-inducible protein 9), a canonical ISG, which is significantly correlated at the protein level with IFNA2, IFNA6 and IFNB1 (**Figure 2 – supplement 2A**). Additionally, IFN ligands often display highly dissimilar, even opposite, relationships to certain proteomics signatures. This is clearly illustrated by the PI3K/AKT/mTOR signature, which was positively correlated with some ligands and negatively correlated with others (**Figure 2E**, compare correlations to HRAS for IFNA1, IFNA6 and IFNB1 in **Figure 2 – supplement 2B**).

To probe further into this phenomenon, we examined the top 5 positively and negatively correlated epitopes for each ligand using unsupervised clustering analysis, which revealed many specialized relationships with potential relevance to COVID19 pathophysiology (**Figure 2 – supplement 2C**). For example, several chemokines involved in immune control showed differential associations, such as CXCL10 (IP10, compare IFNG to IFNW1 in **Figure 2F**); CX3CL1 (fractalkine, compare IFNA10 to IFNA6 in **Figure 2G**); CCL7 (MCP3, compare IFNA2 to IFNA16 in **Figure 2 – supplement 2D**); and CCL5 (RANTES, compare IFNB1 to IFNA10 in **Figure 2 – supplement 2E**). Notably, the top positive correlations for IFNB1 are dominated by proteins stored in alpha granules of platelets, such as PPBP (multiple SOMAscan^®^ aptamers), PDGFA, PDGFD, and PF4 (**Figure 2 – supplement 2C, Supplementary file 8**). These markers of platelet degranulation are also associated, albeit to a lesser degree, with IFNA6, but not so with other ligands (**Figure 2 – supplement 2C**, compare IFNB1 to IFNA10 in **Figure 2H**). This suggests that IFNB1 production is associated with platelet activation, which could be interpreted as a sign of endothelial damage at sites producing IFNB1. A subset of IFN ligands showed strong associations with components of the complement cascade, such as C1QC (**Figure 2 – supplement 2C**, compare IFNA2 to IFNB1 in **Figure 2I**). The top correlated epitope for IFNA10 is TRIL (TRL4 interactor with leucine-rich repeats), a component of the TLR4 complex, but this association was not clear for many other IFN ligands (**Figure 2 – supplement 2C**, compare IFNA10 to IFNA6 in **Figure 2 – supplement 2F**). KIR3DL2 and KIR3DS1, two killer cell immunoglobulin-like receptors (KIRs) expressed by Natural Killer (NK) cells and subtypes of T cells were strongly correlated with a subset of ligands, most prominently IFNA6, but not others (**Figure 2 – supplement 2C**, compare IFNA6 to IFNA10 in **Figure 2 – supplement 2G**). OLFM4 (Olfactomedin 4), a protein selectively expressed in inflamed colonic epithelium (19), was strongly associated with IFNA4/16, but not other ligands (**Figure 2 – supplement 2C**, compare IFNA4/16 versus IFNA10 in **Figure 2 – supplement 2H**). Altogether, these results reveal that circulating levels of different IFN ligands associate with proteomics signatures indicative of multiple pathophysiological processes, such as tissue-specific inflammation, complement activation, and endothelial damage.

### Differential relationship between IFN ligands and seroconversion status

Next, we analyzed correlations between the 12 IFN ligands and the MS plasma proteomics dataset. The MS proteomics platform is highly complementary to the SOMAscan^®^ dataset, as it detects many abundant proteins for which SOMAmer^®^ reagents are not available, such as various immunoglobulins (Igs). Using a linear model adjusting for age and sex, we identified 70 proteins differentially abundant in the plasma of COVID19 patients (**Figure 3 – supplement 1A, Supplementary file 10**). Of the 28 significantly elevated proteins, 17 of them are Igs (labeled green in **Figure 3 – supplement 1A**), potentially indicative of production of anti-SARS-CoV-2 antibodies in COVID19 patients. We then defined Spearman correlations between IFN ligands and all proteins detected by MS (**Figure 3 – supplement 1B, Supplementary file 11**) and visualized the top 5 positively and negatively correlated proteins for each ligand via unsupervised hierarchical clustering (**Figure 3 – supplement 2A**). This analysis confirmed some observations made with the SOMAscan^®^ dataset, but also revealed several new associations. First, a subset of ligands associates strongly with recognizable IFN-inducible proteins such as B2M (beta-2-microglobulin, compare IFNA7/17/21 to IFNA6 in **Figure 3A**), and LGALS3BP (galectin 3 binding protein, compare IFNA4/16 to IFNA6 in **Figure 3 – supplement 2B**). Second, many of the same ligands associate with elevated levels of complement subunits such as C2 (compare IFNL1 to IFNA6 in **Figure 3B**) and C9 (compare IFNA2 to IFNA6 in **Figure 3 – supplement 2C**). Third, several key regulators of coagulation and fibrinolysis were significantly associated with specific ligands. Salient examples include HABP2 (Hyaluronan Binding Protein 2, compare IFNA2 to IFNB1 in **Figure 3C**), FGA (Fibrinogen Alpha Chain, compare IFNL1 to IFNA16 in **Figure 3 – supplement 2D**), F13B (Coagulation Factor XIII B Chain, compare IFNA10 to IFNA6 in Figure S6E), and PROZ (Protein Z, compare IFNW1 to IFNB1 in **Figure 3 – supplement 2F**). Fourth, very distinctly, IFNB1, and to a lesser degree IFNA6, associate positively with markers of platelet degranulation such as PF4, THBS1, PPBP, MMRN1, and SPARC (**Figure 3 – supplement 2A**, compare IFNB1 to IFNA10 in **Figure 3D**). Lastly, IFN ligands have clearly distinct relationships to a subset of immunoglobulin heavy and light chain variable domain peptides, that were either strongly positively or negatively regulated with the levels of specific ligands (compare IFNA2 to IFNA6 in **Figure 3E** and IFNA1 to IFNB1 in **Figure 3F**). This result could potentially be explained by varying levels of IFN ligands upon seroconversion (16).

**Figure 3.**
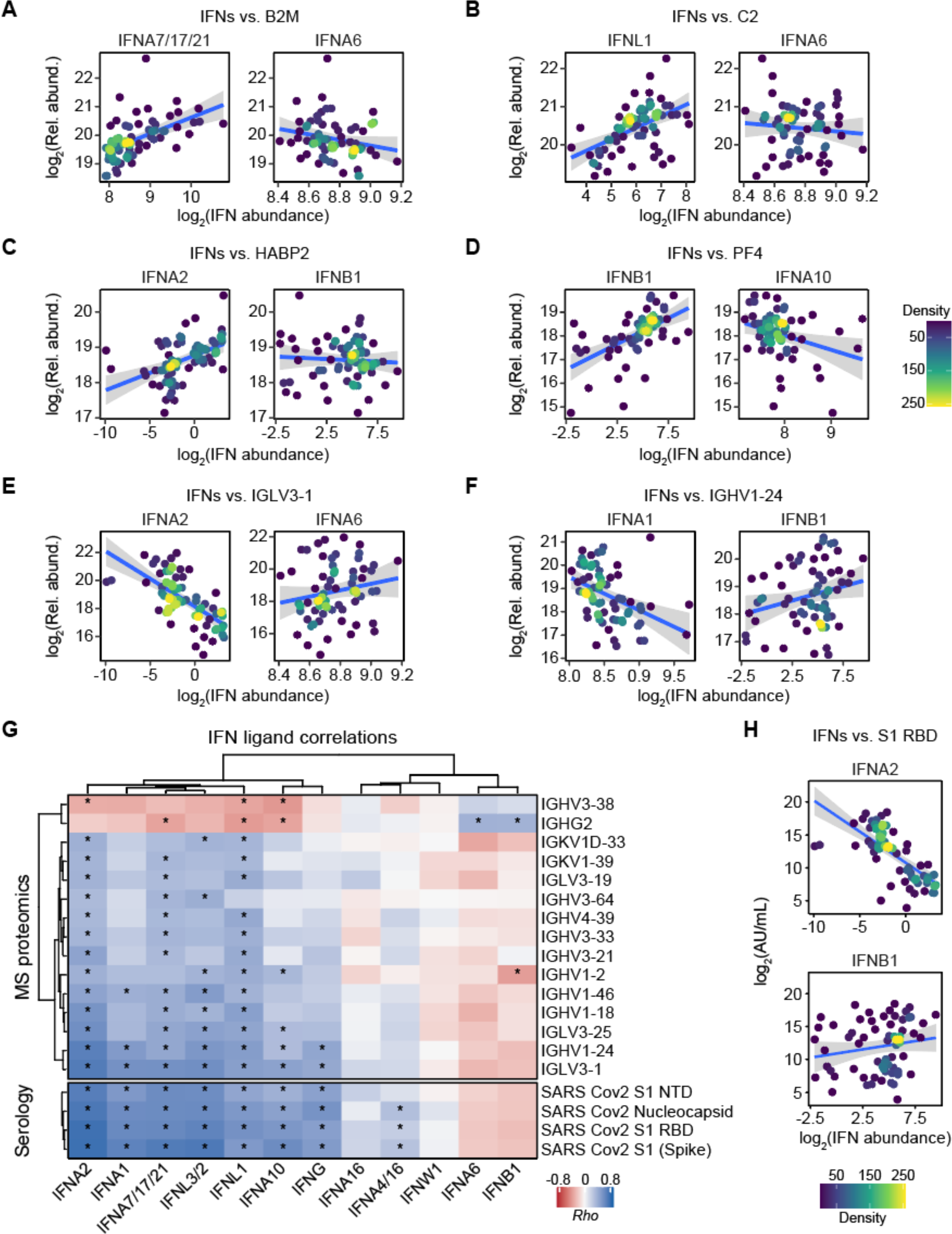
Differential association of IFN ligands with seroconversion. (**A-F**) Scatter plots comparing relationships between plasma proteins, as measured by MS proteomics, and the indicated IFNs in COVID19-positive patients. Points are colored by density; blue lines represent linear model fit with 95% confidence intervals in grey. (**G**) Heatmap representing correlations between IFN ligands and plasma levels of immunoglobulin subunits (top), as measured by MS proteomics, or antibody reactivity against SARS-CoV-2 (bottom), as measured by immunoassays. Only immunoglobulin subunits with at least two significant correlations are shown. Values displayed are Spearman correlation scores (*Rho*); asterisks indicate significant correlations (10% FDR); columns and rows are grouped by hierarchical clustering. (**H**) Scatter plots comparing relationships between plasma antibody reactivity against SARS-CoV-2 S1 RBD region and the indicated IFNs in COVID19-positive patients. Points are colored by density; blue lines represent linear model fit with 95% confidence intervals in grey. See also Figure 3 **– supplement 1** and Figure 3 **– supplement 2**.

In order to investigate in detail the interplay between specific IFN ligands, immunoglobulin expression, and seroconversion, we examined correlations between the ligands and all immunoglobulin variable domains detected by MS proteomics, as well as seroconversion assays used to detect IgGs against SARS-CoV-2 peptides (S1 full length, spike; S1 N-terminus; and S1 receptor binding domain, RBD; nucleocapsid) (**Figure 3G**). This analysis revealed that a subset of IFN ligands is strongly anticorrelated with seroconversion (e.g., compare IFNA2 to IFNB1 in **Figure 3H**) and specific Ig variable domains that have been previously found enriched in the bloodstream of COVID19 patients, such as IGHV1-24 and IGLV3-1 (20, 21). This could be interpreted as early production of some ligands which subsequently declines with seroconversion (e.g., IFNA2, IFNG), followed by later production of other ligands (e.g IFNA6, IFNB1), potentially from sites where SARS-CoV-2 evades humoral neutralization.

Overall, these results further support the notion of differential action of IFN ligands in COVID19 pathophysiology, suggesting a temporal sequence of IFN production prior and after seroconversion.

### Differential immune cell signatures associated with fluctuations in IFN ligand levels

Next, we investigated the relationship between plasma levels of IFN ligands and circulating immune cells analyzed by mass cytometry which measured 38 features per cell event, including surface markers, activation molecules, and transcription factors. First, we employed the unsupervised clustering algorithm PhenoGraph (22) to identify distinct subpopulations of immune cells in the COVIDome mass cytometry dataset, combined with t-stochastic neighbor embedding (t-SNE) dimensionality reduction to aid in visualization (23, 24), resulting in identification of ∼30 clusters, including subpopulations enriched for cell type-specific markers (see Methods, **Figure 4 – supplement 1A-B**). We then identified clusters whose relative frequency among all live cells was significantly associated with varying IFN ligand levels, using beta regression modelling with adjustment for age and sex (**Figure 4A**, **Figure 4 – supplement 1C-D, Supplementary file 12**). This analysis revealed that multiple IFN ligands are significantly associated with increased abundance of clusters enriched for T cells (Clusters 9, 13, 16, and 27, CD3+) and/or NK cells (Cluster 30, 16+ and/or 56+), while also displaying negative associations with clusters enriched for B cells (Clusters 7, 15, and 24, CD19+) (**Figure 4A**, **Figure 4 – supplement 1C-D**). For example, IFNA1 is positively associated with clusters 9 (CD8+ T cells) and 30 (CD56+ NK cells) and negatively associated with cluster 15 (Switched memory B cells) (**Figure 4 – supplement 1E**).

**Figure 4.**
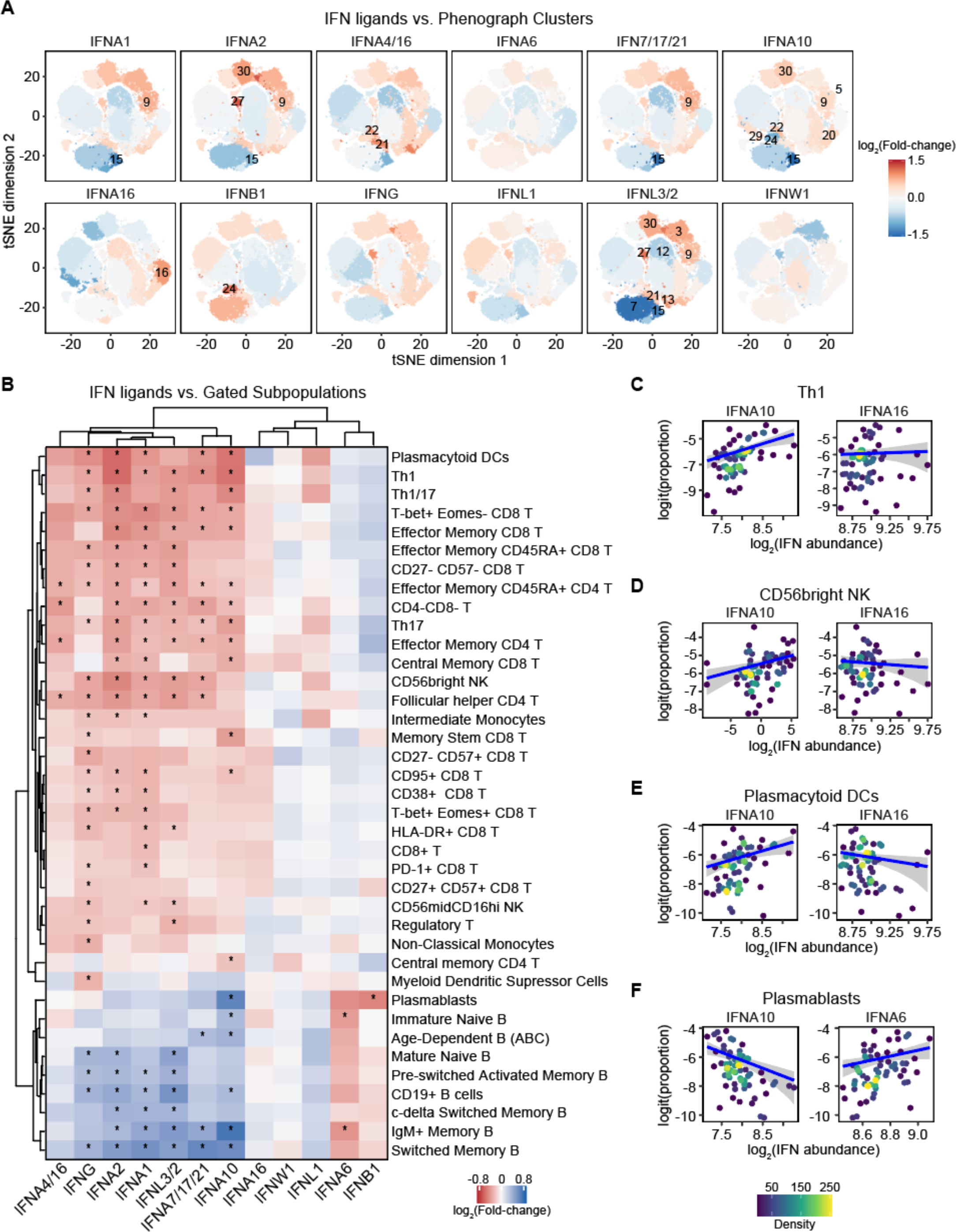
Differential association of IFN ligands with immune cell signatures. (**A**) t-SNE plots of 69,000 cells analyzed by mass cytometry from 69 COVID19-positive patients (1,000 cells each). All cells within each PhenoGraph cluster (as in Figure 4 – supplement 1A) are colored by the fold-change in cluster proportion among live cells per standard deviation of abundance for the indicated IFN, as determined by beta regression analysis, adjusting for Age and Sex; numbers indicate clusters with significant associations with IFN abundance (10% FDR). (**B**) Heatmap representing relationships between IFN ligands and gated subpopulation proportions among live cells, as determined by beta regression analysis. Only subpopulations with at least one significant association are shown. Values displayed are fold-change in cluster proportion among live cells per standard deviation of IFN abundance; asterisks indicate significant associations (10% FDR); columns and rows are grouped by hierarchical clustering. (**C-F**) Scatter plots comparing relationships between gated subpopulation proportions among live cells, and the indicated IFNs in COVID19-positive patients. Points are colored by density; blue lines represent beta regression model fit with 95% confidence intervals in grey. See also Figure 4 **– supplement 1** and Figure 4 **– supplement 2**.

In order to validate and investigate these observations more deeply in relationship to known immune cell sub-populations, we analyzed relations between the IFN ligands and 50+ immune cell types defined by traditional gating based on marker expression (**Figure 4B**, **Figure 4 – supplement 2A, Supplementary file 13,** see **Methods**). This exercise confirmed clear specialized relationships between IFN ligands and specific lymphoid cell subsets. For example, among CD4+ T cells, the T-helper 1 (Th1) subset displays significant positive associations only with IFNA1, IFNA2, IFNA7/17/21, IFNA10, IFNG, and IFNL3/2 (**Figure 4B**, compare IFNA10 to IFNA16 in **Figure 4C**). This pattern was also apparent for many, but certainly not all, T cell subsets (**Figure 4B**). Similarly, NK CD56^bright^ cells also showed differential positive relationships with IFN ligands, with an overall pattern similar to that of key T cell subsets (compare IFNA2 to IFNA16 in **Figure 4D**). Notably, this analysis also revealed significant positive associations between specific ligands and plasmacytoid dendritic cells (pDCs), which are strong producers of IFNs during viral infections (compare IFNA10 to IFNA16 in **Figure 4E**). Many of the IFN ligands positively associated with CD4+ T cell subsets were negatively associated with B cell subsets, while IFNA6 displays the opposite relationship (**Figure 4B**). This is clearly illustrated by plasmablasts (compare IFNA10 versus IFNA6 in **Figure 4F**). These differential associations could be interpreted as a transition from innate T cell-driven responses prior to seroconversion, followed by B cell activation and differentiation toward antibody-producing plasmablasts during seroconversion, along with decreased production of a specific subset of IFN ligands.

Altogether, these results suggest a temporal sequence of IFN ligand production in coordination with changes in the peripheral immune cell compartment, whereby a larger subset of ligands is produced early on during the innate immune response, whereas a few others are associated with development of the adaptive humoral response. An overview of salient IFN ligand associations along the paths of T cell and B cell activation and differentiation is shown in **Figure 4 – supplement 2B**.

### Metabolic signatures of IFN ligand action

Next, we investigated metabolic signatures associated with varying levels of IFN ligands, calculating Spearman correlations for detected metabolites in plasma and red blood cell (RBC) samples against each of the IFN ligands (**Figure 5A-B**, **Figure 5 – supplement 1A-B, Supplementary files 14-15,** see **Methods**). In plasma, significant positive correlations were observed between the tryptophan/indole pathway metabolites kynurenine and 5-hydroxyindoleacetate and IFNG, but not other IFNs (compare IFNG to IFNA16 in **Figure 5C**). In RBCs, kynurenine levels (as well as those of indoxyl, another tryptophan/indole metabolite) showed a strong positive association with IFNG, as well as IFNA7/17/21 (compare IFNG to IFNA7/17/21 in **Figure 5D**). Activation of the kynurenine pathway has been well documented in COVID19 (25-29). Kynurenine production can be stimulated by induction of IDO1 (indoleamine-2,3-dioxygenase 1), an ISG downstream of all three major types of IFN signaling (30).

**Figure 5.**
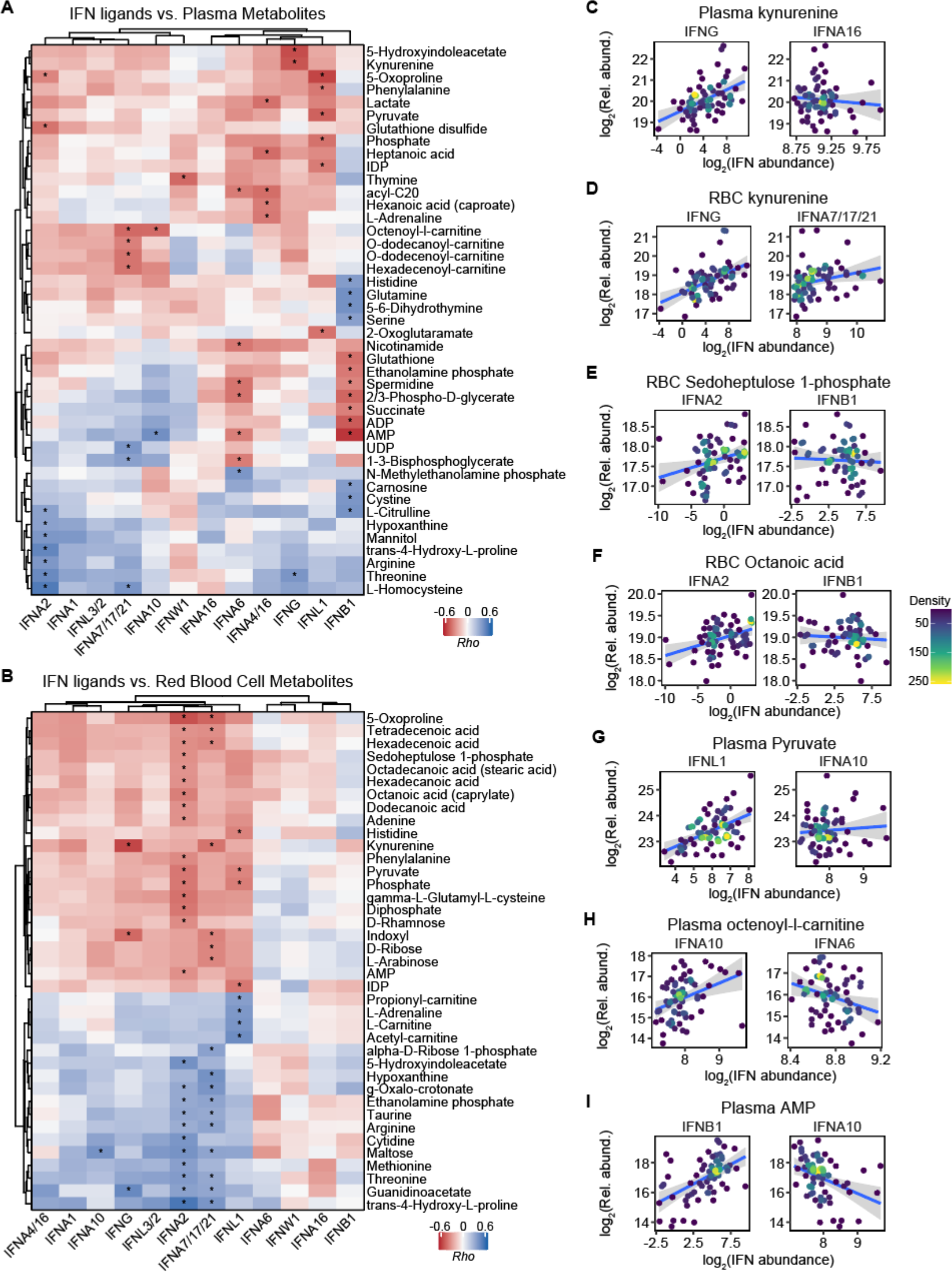
Differential metabolite signatures associated with IFN ligands. (**A-B**) Heatmap representing correlations between IFN ligands and plasma (A) or red blood cell (RBC, B) metabolite levels. Only metabolites with at least one significant correlation are shown. Values displayed are Spearman correlation scores (*Rho*); asterisks indicate significant correlations (10% FDR); columns and rows are grouped by hierarchical clustering. (**C-I**) Scatter plots comparing relationships between select metabolites and the indicated IFNs in COVID19-positive patients. Points are colored by density; blue lines represent linear model fit with 95% confidence intervals in grey. See also Figure 5 **– supplement 1**.

Therefore, it is interesting that this pathway is preferentially associated with IFNG in COVID19. Plasma levels of IFNA2 showed significant positive correlations with the markers of oxidative stress glutathione disulfide and 5-oxoproline, a byproduct of the gamma-glutamyl cycle (**Figure 5A**, compare IFNA2 to IFNW1 in **Figure 5 – supplement 1C**), and negatively associated with markers of endothelial dysfunction and nitric oxide signaling (arginine, citrulline) (**Figure 5A**, compare IFNA2 to IFNW1 in **Figure 5 – supplement 1D**), as well as metabolites of potential bacterial or iatrogenic origin (mannitol) and derived from purine oxidation (hypoxanthine) (**Figure 5A**). In RBCs, IFNA2 had once again strong positive correlations with several markers of oxidative stress (5-oxoproline) or pentose phosphate pathway activation (sedoheptulose phosphate) (**Figure 5B**, compare IFNA2 to IFNB1 in **Figure 5E**), which is required in RBCs to generate reducing equivalent (NADPH) for recycling oxidized glutathione and other NADPH-dependent antioxidant enzymes. IFNA2 levels also positively correlated with fatty acid mobilization in RBCs– perhaps as a result of the activity of peroxiredoxin 6 (31) or phospholipase A2 activity (32, 33) on complex lipids to fuel fatty acid release in the bloodstream to sustain viral capsid formation (34). Of note, among the positive correlates to IFNA2 levels in the fatty acid compartment, we observed only saturated (octanoic, dodecanoic, hexadecenoic, octadecanoic) or monounsaturated fatty acids (tetradecenoic, hexadecenoic, octadecenoic) (compare IFNA2 to IFNB1 in **Figure 5F**), suggestive of limited fatty acid desaturase activation despite the stress induced by the viral infection (35). Several ATP precursors/breakdown products (AMP and adenine) positively correlated with IFNA2 in RBCs, as did pyruvate, phosphate and diphosphate – suggestive of altered glycolysis and overall energetics as a function of IFNA2 signaling. IFNA2 also negatively correlated with several amino acids in RBCs, including the antioxidants taurine, arginine, threonine and methionine – critical for RBC redox damage repair in the face of the incapacity to synthesize new proteins (36).

Plasma IFNL1 significantly correlated with several glycolytic metabolites (e.g. pyruvate, compare IFNL1 to IFNA1 in **Figure 5G**), as well as short chain fatty acids hexanoate and heptanoate, potentially indicative of dysregulation of mitochondrial metabolism in patients with high IFNL1. In RBCs, IFNL1 levels showed positive correlations with the levels of IDP (compare IFNL1 to IFNA6 in **Figure 5 – supplement 1E**) and negative correlations with carnitine and acetyl-carnitine, potentially suggestive of RBC deformability issues (37, 38) as a function of IFNL1 signaling.

Plasma IFNA7 and IFNA10 (and to a lesser extent – IFNA1 and IFNA2) were positively associated with a cluster of acyl-carnitines (including octenoyl, dodecanoyl, dodecenoyl, hexadecenoyl-carnitine, compare IFNA10 to IFNA6 in **Figure 5H**), suggesting an association between these IFN ligands and altered fatty acid oxidation. These data are relevant in light of the role of acyl-carnitines in coagulation (39) and the common thromboembolic complications and dysregulation of coagulation cascades in COVID19 patients associated with inflammatory markers such as IL-6 (40).

Plasma levels of IFNB1 showed strong negative correlation with metabolites tied to the nitric oxide pathway (citrulline), as well as other amine group donors (glutamine, serine) or oxidant stress-related metabolites (carnosine, cystine). On the other hand, IFNB1 positively correlated with the plasma levels of glutathione and spermidine (antioxidant metabolites), succinate (marker of mitochondrial dysfunction), and purinergic agonists involved in vasodilatory/hypoxic responses (ADP and AMP), perhaps produced by hemolytic events (compare IFNB1 to IFNA10 in **Figure 5I** and **Figure 5 – supplement 1F-G**).

Altogether, these results not only confirm metabolic signatures previously associated with IFN signaling (e.g. activation of the kynurenine pathway), but also reveal unexpected associations between specific IFN ligands and diverse metabolic processes with ties to COVID19 pathophysiology.

## DISCUSSION

IFN signaling is a critical component of the innate immune response and a main driver of the antiviral defense. In the context of viral infections, deficiencies in IFN signaling cause profound susceptibility in humans, as demonstrated by various inborn errors of immunity affecting IFN signaling (41). Despite these clear protective effects, dysregulation of IFN signaling can also contribute to diverse pathologies, as exemplified by Type I Interferonopathies, a group of genetic disorders characterized by chronic overproduction of IFN ligands and severe developmental and clinical phenotypes (42). Furthermore, triplication of the four IFN receptors encoded on chromosome 21 (IFNAR1, IFNAR2, IFNGR2, IL10RB) and consequent IFN hyperactivity in individuals with trisomy 21 are thought to contribute to the developmental and clinical hallmarks of Down syndrome, including their high risk of developing severe COVID19 (43-49). In the context of COVID19, the role of IFN signaling has been the subject of much study and debate, with both protective and deleterious effects being documented in different experimental systems and clinical settings (2-4, 10, 14, 15). Within this framework, we provide here a comprehensive analysis of multi-omics signatures associated with production of multiple IFN ligands in hospitalized COVID19 patients, revealing a high degree of diversity and specialization, even among ligands in the same subfamily.

During vertebrate evolution, the IFN ligand gene family has undergone significant expansion through both tandem gene duplication and retrotransposition events, most likely to accommodate increased regulatory diversity and functional specialization (50). Although modest, our current understanding of IFN ligand specialization is increasing. Functional specialization between major Type I, II and III ligands has been revealed by analysis of genetic mutations affecting specific receptors or downstream kinases and transcription factors in both humans and mice (41, 51). For example, it is accepted that deficiencies in IFNG signaling are associated with mycobacterial disease, whereas deficiencies in Type I/III signaling confer susceptibility to viral infections (41). IFN ligand specialization is also evident in the clinical use of recombinant ligands, with IFNB1 being the most effective therapeutic agent for the treatment of multiple sclerosis, whereas IFNA2 preparations are preferred for the treatment of chronic viral infections and some malignancies (52). Despite these advances, little is known about the mechanisms behind these differential effects. In this context, our work provides a valuable resource for future mechanistic research. Although our multi-omics analysis is descriptive in nature and based largely on statistically significant associations that should not be interpreted as cause-effect relationships, its value is confirmed by the many associations observed for which mechanisms have already been established. For example, our unbiased analysis of the transcriptome correlations confirmed that 8 of the 12 ligands tested are indeed significantly and positively associated with a transcriptional program highly enriched for ISGs. Likewise, the association between IFNG and metabolites in the kynurenine pathway can be explained by induction of IDO1, a known ISG, during the inflammatory response elicited by SARS-CoV-2 (25-29). Therefore, using these confirmatory observations as reference points, we propose that the datasets described here will help the field elucidate many novel cause-effect relationships explaining IFN ligand specialization.

The specialized biosignatures of IFN ligand action described here could be due to several non-mutually exclusive mechanisms, such as action through different receptors, differences in affinity or allosteric regulation for the same receptors, as well as differences in the location and timing of ligand production. One limitation of our study is that all measurements were performed from peripheral blood, which can only inform about a subset of the pathophysiological processes modulated by the various ligands. Our study would be highly complemented by studies of IFN ligands in various tissues (e.g. (53)). It is also possible that the specialized biosignatures observed are driven in part by SARS-CoV-2 itself. Like other members of the coronavirus family, SARS-CoV-2 has evolved diverse strategies to evade the antiviral effects of IFN signaling, and it is possible that these escape mechanisms do not affect all IFN ligands equally (54). Despite these limitations, key observations produced by our study include the differential relationship between IFN ligands and the antiviral transcriptional program in circulating immune cells, the specialized relationship between seroconversion, immune cell type abundance and IFN ligand levels, and the distinct metabolic signatures associated with the ligands. Throughout the study, the contrast between IFNA2 and IFNA6 exemplifies these points. Both IFNA2 and IFNA6 are specifically recognized by the reagents employed and significantly upregulated in the COVID19 positive cohort.

However, whereas IFNA2 is strongly associated with the IFN transcriptional program in immune cells, IFNA6 is not. IFNA2 proteomic signatures are enriched for cytokines and chemokines previously linked to IFN signaling, whereas IFNA6 proteomic signatures, similarly to those of IFNB1, are enriched for markers of platelet degranulation. IFNA2 levels decrease with seroconversion, IFNA6 levels do not. Accordingly, IFNA2 abundance associates with increased frequency of various T cell subsets involved in the early antiviral response, while IFNA6 levels correlate with signs of B cell maturation and differentiation. Whereas IFNA2 has the highest number of significant associations in the RBC metabolome of any ligand tested, IFNA6 has none. Therefore, a detailed comparative study of these two IFNA ligands is warranted, including studies in human cell preparations and animal models. In sum, our analyses and datasets provide a rich resource to advance understanding of the IFN ligand family in humans. In order to accelerate the use of these datasets at a global scale, they are made readily available through the COVIDome Explorer Researcher Portal (covidome.org) (17), where users can rapidly recreate the cross-omics correlations described here, investigate any other cross-omics correlations of choice, and download all data for further analysis.

## METHODS

### Study design, participant recruitment, and clinical data capture

Research participants were recruited and consented for participation in the COVID Biobank of the University of Colorado Anschutz Medical Campus [Colorado Multiple Institutional Review Board (COMIRB) Protocol # 20-0685]. Data was generated from deidentified biospecimens and linked to demographics and clinical metadata procured through the Health Data Compass of the University of Colorado under COMIRB Protocol # 20-1700. Participants were hospitalized either at Children’s Hospital Colorado or the University of Colorado Hospital. COVID status was defined by a positive PCR result and/or antibody test within 14 days of the research blood draw. The control cohort consisted of COVID19-negative research participants receiving medical care for a range of conditions, none of them in critical condition at the time of the research blood draw. Cohort characteristics can be found in **Supplementary file 1**.

### Blood processing

Blood samples were collected into EDTA tubes, sodium heparin tubes, and PAXgene Blood RNA Tubes (PreAnalytiX/Qiagen). After centrifugation, EDTA plasma was used for MS proteomics, SOMAscan^®^ proteomics, as well as multiplex immunoassays using MSD technology for both cytokine profiles and seroconversion assays. From sodium heparin tubes, PBMCs were obtained by the Ficoll gradient method before cryopreservation and assembly of batches for MC analysis (see below).

### Whole-blood RNA library preparation and sequencing

RNA was purified from PAXgene Blood RNA Tubes (PreAnalytiX/Qiagen) using a PAXgene Blood RNA Kit (Qiagen), according to the manufacturer’s instructions. RNA quality was assessed using an Agilent 2200 TapeStation and quantified by Qubit (Life Technologies). Globin RNA depletion, poly-A(+) RNA enrichment, and strand-specific library preparation were carried out using a Universal Plus mRNA-Seq with NuQuant, Human Globin AnyDeplete (Tecan). Paired-end 150 bp sequencing was carried out on an Illumina NovaSeq 6000 instrument by the Genomics Shared Resource at the University of Colorado Anschutz Medical Campus.

### Plasma proteomics by SOMAscan^®^ assays

125 μL EDTA plasma was analyzed by SOMAscan^®^ assays using previously established protocols (55). Briefly, each of the 4000+ SOMAmer reagents binds a target peptide and is quantified on a custom Agilent hybridization chip. Normalization and calibration were performed according to SOMAscan^®^ Data Standardization and File Specification Technical Note (SSM-020) (55). The output of the SOMAscan^®^ assay is reported in relative fluorescent units (RFU). Validation of IFN detection was carried out by spiking recombinant human IFN ligands into separate aliquots of a pooled plasma reference sample (10 pg/µL). Data were processed as above and then to account for background signal in the reference sample, the median relative abundance measured by each SOMAscan^®^ aptamer reagent across all samples was subtracted from the corresponding values for each spike-in sample. Recombinant human IFN ligands were obtained from PBL Assay Science (Piscataway, NJ 08854 USA), with the following catalog numbers: 11002-1 (Human Interferon Alpha Sampler Set: IFNA1, IFNA2, IFNA4, IFNA5, IFNA6, IFNA7, IFNA8, IFNA10, IFNA14, IFNA16, IFNA17, IFNA21); 11725-1 (IFNL1); 11720-1 (IFNL2); 11730-1 (IFNL3); 11500-1 (IFNG); 11420-1 (IFNB1).

### Cytokine profiling and seroconversion by multiplex immunoassay

Multiplex immunoassays MSD assays were performed on EDTA plasma aliquots following manufacturer’s instructions (Meso Scale Discovery, MSD). *Absolute* values were obtained by extrapolation against a standard curve using provided calibrators. Validation of IFN detection was carried out by spiking a range of concentrations of recombinant IFN ligands into separate aliquots of a pooled plasma reference sample followed by measurement as above. Recombinant human IFN ligands were obtained from PBL Assay Science (Piscataway, NJ 08854 USA), as described above. Seroconversion assays against SARS-CoV-2 proteins were performed in a multiplex immunoassay using the IgG detection readout according to manufacturer’s instructions (MSD). *Relative* values were obtained by extrapolation against a standardized curve consisting of pooled COVID19-positive reference plasma (56).

### Plasma proteomics by mass spectrometry

Plasma samples were digested in S-Trap filters (Protifi, Huntington, NY) according to the manufacturer’s procedure. Briefly, a dried protein pellet prepared from organic extraction of patient plasma was solubilized in 400 µl of 5% (w/v) SDS. Samples were reduced with 10 mM DTT at 55°C for 30 min, cooled to room temperature, and then alkylated with 25 mM iodoacetamide in the dark for 30 min. Next, a final concentration of 1.2% phosphoric acid and then six volumes of binding buffer [90% methanol; 100 mM triethylammonium bicarbonate (TEAB); pH 7.1] were added to each sample. After gentle mixing, the protein solution was loaded into an S-Trap filter, spun at 2000 rpm for 1 min, and the flow-through collected and reloaded onto the filter. This step was repeated three times, and then the filter was washed with 200 μL of binding buffer 3 times. Finally, 1 μg of sequencing-grade trypsin (Promega) and 150 μL of digestion buffer (50 mM TEAB) were added onto the filter and digestion carried out at 47 °C for 1 h. To elute peptides, three stepwise buffers were applied, 200 μL of each with one more repeat, including 50 mM TEAB, 0.2% formic acid in H_2_O, and 50% acetonitrile and 0.2% formic acid in H_2_O. The peptide solutions were pooled, lyophilized and resuspended in 1 mL of 0.1 % FA. 20 µl of each sample was loaded onto individual Evotips for desalting and then washed with 20 μL 0.1% FA followed by the addition of 100 μL storage solvent (0.1% FA) to keep the Evotips wet until analysis. The Evosep One system (Evosep, Odense, Denmark) was used to separate peptides on a Pepsep column, (150 µm internal diameter, 15 cm) packed with ReproSil C18 1.9 µm, 120A resin. The system was coupled to a timsTOF Pro mass spectrometer (Bruker Daltonics, Bremen, Germany) via a nano-electrospray ion source (Captive Spray, Bruker Daltonics). The mass spectrometer was operated in PASEF mode. The ramp time was set to 100 ms and 10 PASEF MS/MS scans per topN acquisition cycle were acquired. MS and MS/MS spectra were recorded from m/z 100 to 1700. The ion mobility was scanned from 0.7 to 1.50 Vs/cm^2^. Precursors for data-dependent acquisition were isolated within ± 1 Th and fragmented with an ion mobility-dependent collision energy, which was linearly increased from 20 to 59 eV in positive mode. Low-abundance precursor ions with an intensity above a threshold of 500 counts but below a target value of 20000 counts were repeatedly scheduled and otherwise dynamically excluded for 0.4 min. Raw data file conversion to peak lists in the MGF format, downstream identification, validation, filtering and quantification were managed using FragPipe version 13.0. MSFragger version 3.0 was used for database searches against a Human isoform-containing UniProt fasta file (version 08/11/2020) with decoys and common contaminants added. The identification settings were as follows: Trypsin, Specific, with a maximum of 2 missed cleavages, up to 2 isotope errors in precursor selection allowed for, 10.0 ppm as MS1 and 20.0 ppm as MS2 tolerances; fixed modifications: Carbamidomethylation of C (+57.021464 Da), variable modifications: Oxidation of M (+15.994915 Da), Acetylation of protein N-term (+42.010565 Da), Pyrolidone from peptide N-term Q or C (-17.026549 Da). The Philosopher toolkit version 3.2.9 (build 1593192429) was used for filtering of results at the peptide and protein level at 0.01 FDR. Label-free quantification was performed by AUC integration with matching between all runs using IonQuant.

### Mass cytometry analysis of immune cell types

Cryopreserved PBMCs were thawed, washed twice with Cell Staining Buffer (CSB) (Fluidigm), and counted with an automated cell counter (Countess II - Thermo Fisher Scientific). Extracellular staining on live cells was done in CSB for 30 min at room temperature, in 3-5^10^6^ cells per sample. Cells were washed with 1X PBS (Fluidigm) and stained with 1 mL of 0.25 mM cisplatin (Fluidigm) for 1 min at room temperature for exclusion of dead cells. Samples were then washed with CSB and incubated with 1.6% PFA (Electron Microscopy Sciences) during 10 min at room temperature. Samples were washed with CBS and barcoded using a Cell-IDTM 20-Plex Pd Barcoding Kit (Fluidigm) of lanthanide-tagged cell reactive metal chelators that will covalently label samples with a unique combination of palladium isotopes, then combined. Surface staining with antibodies that work on fixed epitopes was performed in CSB for 30 min at room temperature (see **Supplementary file 16** for antibody information). Cells were washed twice with CSB and fixed in Fix/Perm buffer (eBioscience) for 30 min, washed in permeabilization buffer (eBioscience) twice, then intracellular factors were stained in permeabilization buffer for 45 min at 4°C. Cells were washed twice with Fix/Perm Buffer and were labeled overnight at 4°C with Cell-ID Intercalator-Ir (Fluidigm) for DNA staining. Cells were then analyzed on a Helios instrument (Fluidigm). To make all samples comparable, pre-processing of mass cytometry data included normalization within and between batches via polystyrene beads embedded with lanthanides as previously described (57). Files were debarcoded using the Matlab DebarcoderTool (58). Then normalization again between batches relative to a reference batch based on technical replicates (59).

### Mass spectrometry-based metabolomics of plasma and red blood cells

#### Sample extraction

Samples were thawed on ice and extracted via a modified Folch method (chloroform/methanol/water 8:4:3), which completely inactivates other coronaviruses, such as MERS-CoV. Briefly, 20 μL of sample was diluted in 130 μL of LC-MS grade water, 600 μL of ice-cold chloroform/methanol (2:1) was added, and the samples were vortexed for 10 seconds. Samples were then incubated at 4°C for 5 minutes, quickly vortexed (5 seconds), and centrifuged at 14,000 *g* for 10 minutes at 4°C. The top (i.e., aqueous) phase was transferred to a new tube for metabolomics analysis and flash frozen. The bottom (i.e., organic) phase was transferred to a new tube for lipidomics analysis, then dried under N_2_ flow.

#### UHPLC-MS metabolomics

Analyses were performed using a Vanquish UHPLC coupled online to a Q Exactive high resolution mass spectrometer (Thermo Fisher Scientific, Bremen, Germany). Samples (10 uL per injection) were randomized and analyzed in positive and negative electrospray ionization modes (separate runs) using a 5-minute C18 gradient on a Kinetex C18 column (Phenomenex) as described (60). Data were analyzed using Maven (Princeton University, Princeton, NJ, USA) in conjunction with the KEGG database and an in-house standard library.

### Biostatistics and bioinformatics analyses

Preprocessing, statistical analysis, and plot generation for all datasets was carried out using R (R 4.0.1 / Rstudio 1.3.959 / Bioconductor v 3.11) (61-63), as detailed below.

#### Analysis of transcriptome data

RNA-seq data yield was ∼40-80 x 10^6^ raw reads and ∼32-71 x 10^6^ final mapped reads per sample. Reads were demultiplexed and converted to fastq format using bcl2fastq (bcl2fastq v2.20.0.422). Data quality was assessed using FASTQC (v0.11.5) (https://www.bioinformatics.babraham.ac.uk/projects/fastqc/) and FastQ Screen (v0.11.0, https://www.bioinformatics.babraham.ac.uk/projects/fastq_screen/). Trimming and filtering of low-quality reads was performed using bbduk from BBTools (v37.99)(64) and fastq-mcf from ea-utils (v1.05, https://expressionanalysis.github.io/ea-utils/). Alignment to the human reference genome (GRCh38) was carried out using HISAT2 (v2.1.0)(65) in paired, spliced-alignment mode with a GRCh38 index with a Gencode v33 annotation GTF, and alignments were sorted and filtered for mapping quality (MAPQ > 10) using Samtools (v1.5)**(66)**. Gene-level count data were quantified using HTSeq-count (v0.6.1)(67) with the following options (--stranded=reverse –minaqual=10 –type=exon --mode=intersection-nonempty) using a Gencode v33 GTF annotation file. Differential gene expression in COVID+ versus COVID-was evaluated using DESeq2 (version 1.28.1)(68) in R (version 4.0.1), using q < 0.1 (FDR < 10%) as the threshold for differentially expressed genes.

#### Analysis of SOMAscan^®^ data

Normalized data (RFU) was imported and converted from a SOMAscan^®^ .adat file using a custom R package (SomaDataIO v3.1.0, https://github.com/SomaLogic/SomaDataIO) for use in all subsequent analysis.

#### Analysis of MSD cytokine profiling data

Plasma concentration values (pg/mL) for each of the cytokines and related immune factors measured across multiple MSD assay plates was imported to R, combined, and analytes with >10% of values outside of detection or fit curve range flagged. For each analyte, missing values were replaced with either the minimum (if below fit curve range) or maximum (if above fit curve range) calculated concentration and means of duplicate wells used in all further analysis.

#### Analysis of MS-proteomic data

Raw Razor intensity data were filtered for high abundance proteins by removing those with >70% zero values in both COVID19-negative and COVID19-positive groups. For the remaining 407 abundant proteins, 0 values (8,363 missing values of 44,363 total measurements) were replaced with a random value sampled from between 0 and 0.5x the minimum non-zero intensity value for that protein. Data was then normalized using a scaling factor derived from the global median intensity value across all proteins / sample median intensity across all proteins (69).

#### Analysis of mass cytometry data

MC data was exported as individual FCS files. Within the cytofkit package graphical user interface (v1.11.3) (70), FCS files were imported to R (v4.0.3) using the read.FCS() function from the flowCore package (v2.2.0) (71) and raw intensity values inverse hyperbolic sine transformed using the cytofAsinh() function with cofactor = 5 from the cytofkit package, and 1000 cells per FCS file sampled without replacement for downstream analysis. For visualization, dimensionality reduction was performed using the t-distributed stochastic neighbor embedding (t-SNE) method from the Rtsne package (v0.15) (72), using all markers. Unsupervised clustering, using all markers, was performed using the cytofkit implementation of the PhenoGraph algorithm (22).

Transformed marker expression values for each clustered cell/event were exported and Z-scores calculated across all events for visualization on t-SNE plots. Relative frequencies for each cluster were calculated as proportions of live cells per sample for use in subsequent analyses. For traditionally gated cell subpopulations (gating strategy is described in (16)), relative frequencies were exported from CellEngine as percentages of various parental lineages for use in subsequent analyses.

#### Analysis of LCMS-metabolomics data

Peak intensity data was imported to R. Across the 171 metabolites, 0 values (486 missing values of 21,033 total measurements) were replaced with a random value sampled from between 0 and 0.5x the minimum non-zero intensity value for that metabolite. For downstream analysis, data was then normalized using a scaling factor derived by dividing the global median intensity value across all proteins by each sample median intensity. Median normalization was chosen as it is simple to employ, relies on few assumptions, and performs on-par with more complex normalization techniques, such as linear regression, local regression, total intensity, average intensity, and quantile normalization, in reducing intragroup variation (73), and is one of the non-reference-based normalization methods employed in the widely-used MetaboAnalyst pre-processing module (74).

#### Gene Set Enrichment Analysis (GSEA)

GSEA (75) was carried out using the fgsea package (v 1.14.0) (76) in R (version 4.0.1), using Hallmark gene sets (18) and either log2-transformed fold-changes (for RNA-seq and Somascan) or Spearman *rho* values (for IFN correlations) as the ranking metric. *Interferon Alpha/Gamma Scores*. To capture interferon signaling in each sample as a single value we calculated RNA-seq- or Somascan-based ‘Interferon Alpha’ and ‘Interferon Gamma’ scores as follows: Firstly, Z-scores were calculated from the age- and sex-adjusted concentration values for each gene/protein in each sample, based on the mean and standard deviation of COVID19-negative samples. Secondly, per-sample scores were calculated as the sum of Z-scores for genes/proteins in the Hallmark Interferon Alpha or Hallmark Interferon Gamma Response gene sets (18), filtered to genes/proteins with significant increases in the COVID-positive group (see next section).

#### Differential abundance analysis

For RNAseq, gene-level differential expression in COVID+ versus COVID- was evaluated using DESeq2 (version 1.28.1)(68) in R (version 4.0.1), with q < 0.1 (FDR < 10%) as the threshold for differentially expressed genes, and considering only genes with ≥ 0.5 counts-per-million in at least two samples. Differential abundance analysis for SOMAscan^®^ proteomics, MSD cytokine profiling, MS proteomics, and LCMS metabolomics was performed using linear models in R (version 4.0.1) with log2 concentration/abundance as the outcome/dependent variable and COVID19 status as the predictor/independent variable, with adjustment for Age and Sex. Multiple hypothesis correction was performed with the Benjamini-Hochberg method using a false discovery rate (FDR) threshold of 10% (q<0.1).

#### Correlation analysis

To identify features in each dataset that correlate with plasma levels of the 12 IFN ligands in COVID19 positive samples, Spearman *rho* values and p-values were calculated against the Sex/Age-adjusted values for each dataset using the *rcorr* function from the Hmisc package (v 4.4-0) (77), with Benjamini-Hochberg correction of p-values and an estimated FDR threshold of 0.1. For visualization, Heatmaps and XY scatter plots, with points colored by local density using a custom density function, were generated using the ComplexHeatmap (v2.4.2) (78) and the ggplot2 (v3.3.1) (79) packages. Extreme outlier data points (above Q3 + 3xIQR or below Q1 – 3XIQR) were removed.

#### Beta regression analysis of MC data

To identify cell clusters or gated cell subsets for which relative frequencies are associated with plasma levels of the 12 IFN ligands in COVID19 positive samples, beta regression analysis was carried out using the betareg package (v3.1-4) (80), with each model using cell cluster/subset proportions (relative frequency) as the outcome/dependent variable and log2-transformed IFN abundance values as the independent/predictor variable, with adjustment for Age and Sex, and a logit link function. Effect sizes (as fold-change per unit IFN abundance) for each IFN ligand were obtained by exponentiation of beta regression model coefficients. For comparison across IFN ligands as in volcano plots and heatmaps, beta regression model coefficients were multiplied by the standard deviation of the corresponding ligand before exponentiating to give ‘standardized’ fold-changes per standard deviation of IFN abundance. Standardized fold-changes from each model were visualized by overlaying on t-SNE plots or as heatmaps using the ggplot2 (v3.3.1) (79) and ComplexHeatmap (v2.4.2) (78) packages. For visualization of individual IFN ligand vs. cluster/subset examples, data points were visualized as XY scatter plots, with points colored by local density using a custom function, and overlaid with beta regression fit curves and 95% confidence intervals extracted from model objects using the *ggemmeans()* function from the ggeffects package (v1.1.0) (81).

## DATA AVAILABILITY STATEMENT

Data used in this manuscript are available via the COVIDome Explorer online researcher portal (covidome.org), as described in (17). The RNAseq data have been deposited in NCBI Gene Expression Omnibus, with series accession number GSE167000. The SOMAscan® Proteomics, MSD Cytokine Profiles, and Sample Metadata files have been deposited in Mendeley under entry doi:10.17632/2mc6rrc5j3.2. The mass spectrometry proteomics data have been deposited to the ProteomeXchange Consortium via the PRIDE partner repository (75) with the dataset identifier PXD022817. The mass cytometry data has been deposited in Flow Repository (flowrepository.org) (76) with Repository ID FR-FCM-Z367. Raw metabolomics MS data files and annotated reports are available at the NIH Common Fund’s National Metabolomics Data Repository (NMDR) website (supported by NIH grant U2C-DK119886), the Metabolomics Workbench (metabolomicsworkbench.org) under the Project ID PR001110. These data can be accessed directly via the Project DOI: 10.21228/M8739H.

## Data Availability

Data used in this manuscript are available via the COVIDome Explorer online researcher portal (covidome.org). The RNAseq data have been deposited in NCBI Gene Expression Omnibus, with series accession number GSE167000. The SOMAscan® Proteomics, MSD Cytokine Profiles, and Sample Metadata files have been deposited in Mendeley under entry doi:10.17632/2mc6rrc5j3.2. The mass spectrometry proteomics data have been deposited to the ProteomeXchange Consortium via the PRIDE partner repository with the dataset identifier PXD022817. The mass cytometry data has been deposited in Flow Repository (flowrepository.org) with Repository ID FR-FCM-Z367. Raw metabolomics MS data files and annotated reports are available at the NIH Common Fund's National Metabolomics Data Repository (NMDR) website (supported by NIH grant U2C-DK119886), the Metabolomics Workbench (metabolomicsworkbench.org) under the Project ID PR001110. These data can be accessed directly via the Project DOI: 10.21228/M8739H.

https://www.metabolomicsworkbench.org/data/DRCCMetadata.php?Mode=Project&ProjectID=PR001110

https://flowrepository.org/id/RvFrSYioKeUdYHXdkTD9TQPAXt4PqdkB5eie82h11JgAGSCQIneLKpcKd81Nzgwq

http://proteomecentral.proteomexchange.org/cgi/GetDataset?ID=PXD022817

https://data.mendeley.com/datasets/2mc6rrc5j3/1

https://www.ncbi.nlm.nih.gov/geo/query/acc.cgi?acc=GSE167000

## AKNOWLEDGMENTS

We are grateful to Dr. Thomas Flaig and the Office of the Vice Chancellor for Research (OVCR) at the University of Colorado Anschutz Medical Campus for their leadership in setting up the COVID19 Biobank at the University of Colorado, and also to the COVID19 Biobank Steering Committee for overall support of this project. We thank members of the Biorepository Shared Resource, especially Dr. Adrie Van Bokhoven, Zachary Grasmick, and Hannah Schumman; members of the Human Immune Monitoring Shared Resource, especially Dr. Jill Slansky, Jodi Livesay, Troy Schedin, and Jennifer McWilliams; members of the Flow Cytometry Shared Resource of the University of Colorado Cancer Center, specially Eric Cambley, Alistair Acosta, Christine Childs, and Kristina Terrell; as well as Aaron Issaian for assistance with MS proteomics data analysis. We also thank the SomaLogic team for their support and the Meso Scale Discovery team for generous support with seroconversion assays. We are grateful to Dr. Ian Brooks, Michelle Edelmann and the rest of the Health Data Compass team for the clinical data.

## AUTHOR CONTRIBUTIONS

Conceptualization: MDG, JME, KDS, AAM, TDB, AD, KCH, KRJ, EWYH Investigation: PA, KPS, REG, RB, KRJ, MD, JAR, FG, FC, TG, AD, KCH

Data curation: MDG, SR, KTK, JRS, TG, RB Software: MDG, KTK, JRS, TG, RB

Formal Analysis: MDG, KTK, JME, JRS, PA, TG, RB Visualization: MDG, JME, PA

Funding acquisition: JME, TDB, KCH, AD, KDS, EWYH Writing – original draft: JME, MDG

Writing – review & editing: all authors.

## SUPPLEMENTARY FILES LEGENDS

**Supplementary file 1. Cohort characteristics.** Table summarizing cohort characteristics. Information pertaining less than 10% of the cohort is indicated as <10% to prevent potential reidentification.

**Supplementary file 2. Transcriptome differential expression by COVID status.** Results of DESeq2 differential expression analysis of whole blood RNA-seq data in COVID-19-positive vs. -negative samples.

**Supplementary file 3. GSEA of transcriptome by COVID status.** Results from Gene Set Enrichment Analysis (GSEA) of Hallmark gene sets using RNA-seq fold-change COVID-19-positive vs. negative as the ranking metric.

**Supplementary file 4. IFN ligands vs transcriptome correlations.** Results of Spearman correlation analysis between plasma levels of IFN ligands and whole blood RNA-seq gene-level expression in COVID-19-positive samples.

**Supplementary file 5. GSEA of IFN ligands vs transcriptome correlations.** Results from Gene Set Enrichment Analysis (GSEA) of Hallmark gene sets using Spearman correlation scores for IFN ligands vs. whole blood RNA-seq gene-level expression as the ranking metric.

**Supplementary file 6. SOMAscan^®^ proteomics differential abundance by COVID status.** Results of linear model differential abundance analysis of plasma SOMAscan^®^ proteomics data in COVID-19-positive vs. -negative samples.

**Supplementary file 7. GSEA of SOMAscan^®^ proteomics by COVID status.** Results from Gene Set Enrichment Analysis (GSEA) of Hallmark gene sets using SOMAscan^®^ proteomics fold-change COVID-19-positive vs. negative as the ranking metric.

**Supplementary file 8. IFN ligands vs SOMAscan^®^ proteomics correlations.** Results of Spearman correlation analysis between plasma levels of IFN ligands and SOMAscan^®^ proteomics data in COVID-19-positive samples.

**Supplementary file 9. GSEA of IFN ligands vs SOMAscan proteomics correlations.** Results from Gene Set Enrichment Analysis (GSEA) of Hallmark gene sets using Spearman correlation scores for IFN ligands vs. SOMAscan^®^ proteomics data as the ranking metric.

**Supplementary file 10. MS proteomics differential abundance by COVID status.** Results of linear model differential abundance analysis of plasma mass spectrometry (MS) proteomics data in COVID-19-positive vs. -negative samples.

**Supplementary file 11. IFN ligands vs MS proteomics correlations.** Results of Spearman correlation analysis between plasma levels of IFN ligands and mass spectrometry (MS) proteomics data in COVID-19-positive samples.

**Supplementary file 12. IFN ligands vs mass cytometry clusters beta regression.** Results of beta regression analysis of relative frequency data for PhenoGraph-defined subpopulation clusters against plasma levels of IFN ligands in COVID-19-positive samples.

**Supplementary file 13. IFN ligands vs mass cytometry gated subpopulations beta regression.** Results of beta regression analysis of relative frequency data for cell subpopulations defined by manual gating against plasma levels of IFN ligands in COVID-19-positive samples.

**Supplementary file 14. IFN ligands vs plasma metabolomics correlations.** Results of Spearman correlation analysis between plasma levels of IFN ligands and plasma metabolomics data in COVID-19-positive samples.

**Supplementary file 15. IFN ligands vs RBC metabolomics correlations.** Results of Spearman correlation analysis between plasma levels of IFN ligands and red blood cell (RBC) metabolomics data in COVID-19-positive samples.

**Supplementary file 16. Antibodies used in mass cytometry.** List of antibodies used in mass cytometry. Column A indicates the antibody target, column B indicates the element conjugated to the antibody, column C indicates the mass of the element, column D indicates the manufacturer, column E indicates the catalog number, column F indicates the clone number, and column G indicates the type of stain protocol used (fixed, live or fixed with permeabilization).

---

**Figure 1 – supplement 1.**
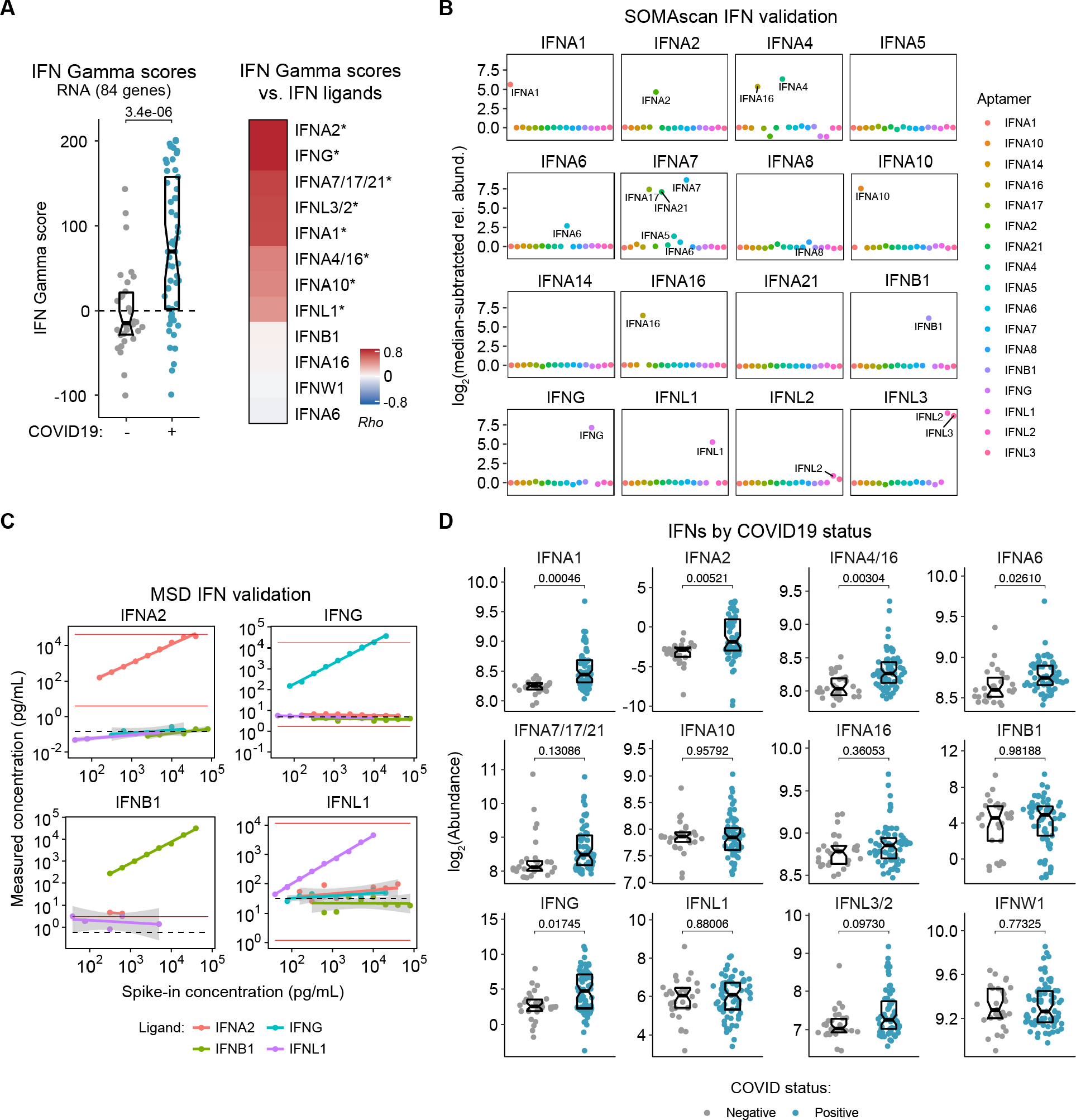
Whole-blood RNA-based IFN Gamma scores and validation of plasma IFN ligand measurements. (**A**) Sina plot of RNA-based IFN Gamma scores, separated by COVID19 status and ranked heatmap representing correlations between RNA-based IFN Gamma scores and plasma levels of each IFN ligand. IFN Gamma scores were calculated for each research participant by summing Z-scores for 84 differentially expressed genes from the IFN Gamma response Hallmark gene set from MSigDB. Z-scores were calculated from the adjusted concentration values for each gene in each sample, based on the mean and standard deviation of COVID19-negative samples. Data are presented as a modified sina plot with box indicating median and interquartile range. Heatmap values displayed are Spearman correlation coefficients (Rho); asterisks indicate significant correlations (10% FDR). (**B**) Validation of IFN ligand detection by SOMAscan^®^ assay. Each plot represents relative abundance above background measured by IFN-targeting SOMAscan^®^ aptamers (indicated by color) for each recombinant IFN ligand spike-in (indicated by plot labels). (**C**) Validation of IFN ligand detection by MSD immunoassay. Plots show the relationship between measured concentration and spike-in concentration for each recombinant IFN ligand (indicated by point and line color) for each assay (indicated by plot labels). Horizontal dashed lines indicate measured concentrations for the pooled plasma sample with no spike-in; red lines indicate manufacturer-stated detection limits. (**D**) Sina plots comparing abundance for the indicated IFNs in COVID19-negative (-) vs. -positive (+) plasma samples. Data are presented as modified sina plots with boxes indicating median and interquartile range. Numbers above brackets are q-values for Mann–Whitney tests.

**Figure 1 – supplement 2.**
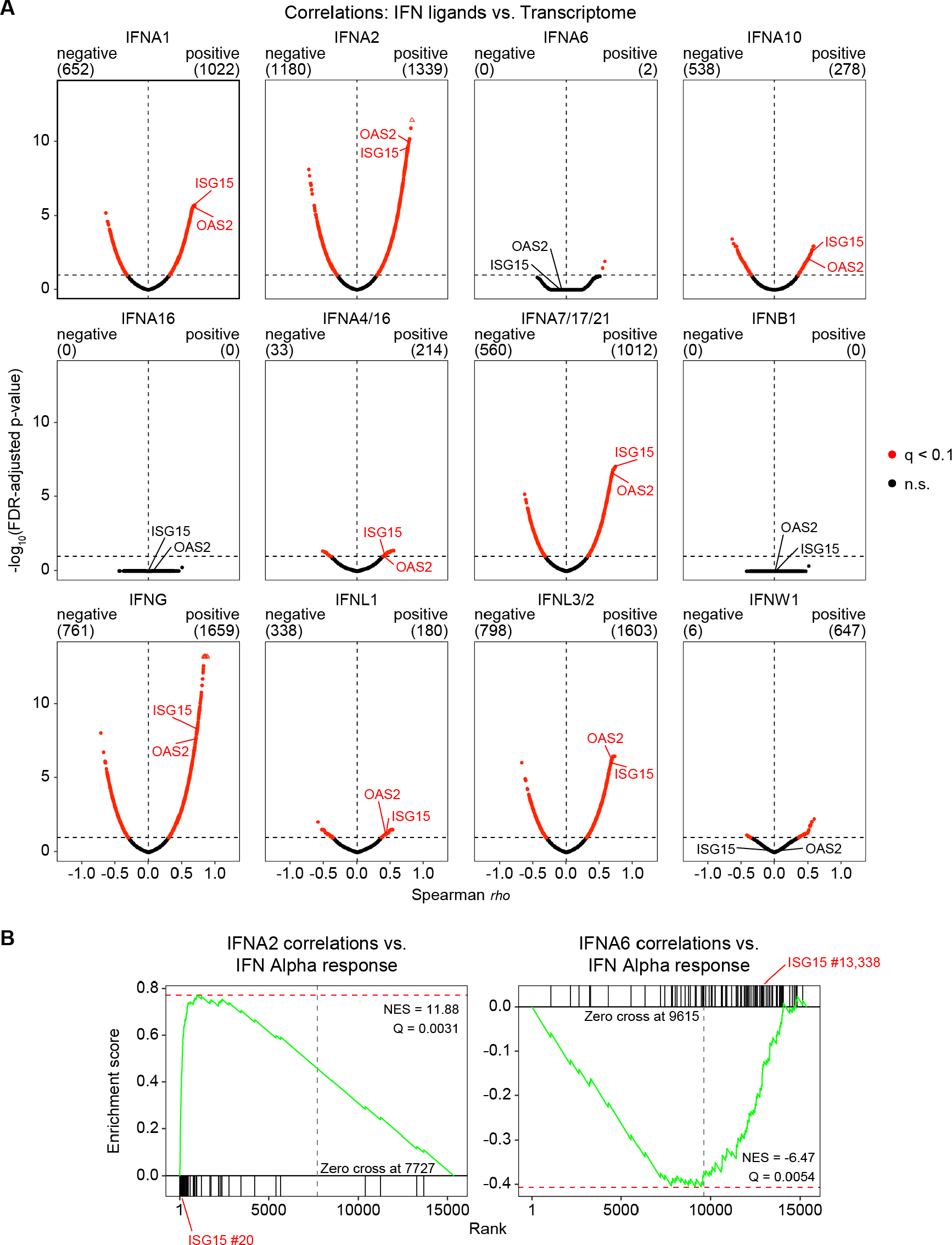
Correlation analysis and GSEA for IFN ligands vs. whole blood transcriptome. (**A**) Volcano plots for Spearman correlation analysis of IFN ligands vs. gene-level RPKM values. Horizontal dashed line indicates an FDR threshold of 10% (q < 0.1); red points and numbers above plots indicate significant genes at this threshold. (**B**) Gene set enrichment analysis (GSEA) plots for the IFN Alpha Response Hallmark gene set from MSigDB. Green lines indicate cumulative enrichment score; black bars indicate gene set hits among all genes ranked by log2(fold change) for COVID19-positive vs. -negative samples.

**Figure 2 – supplement 1.**
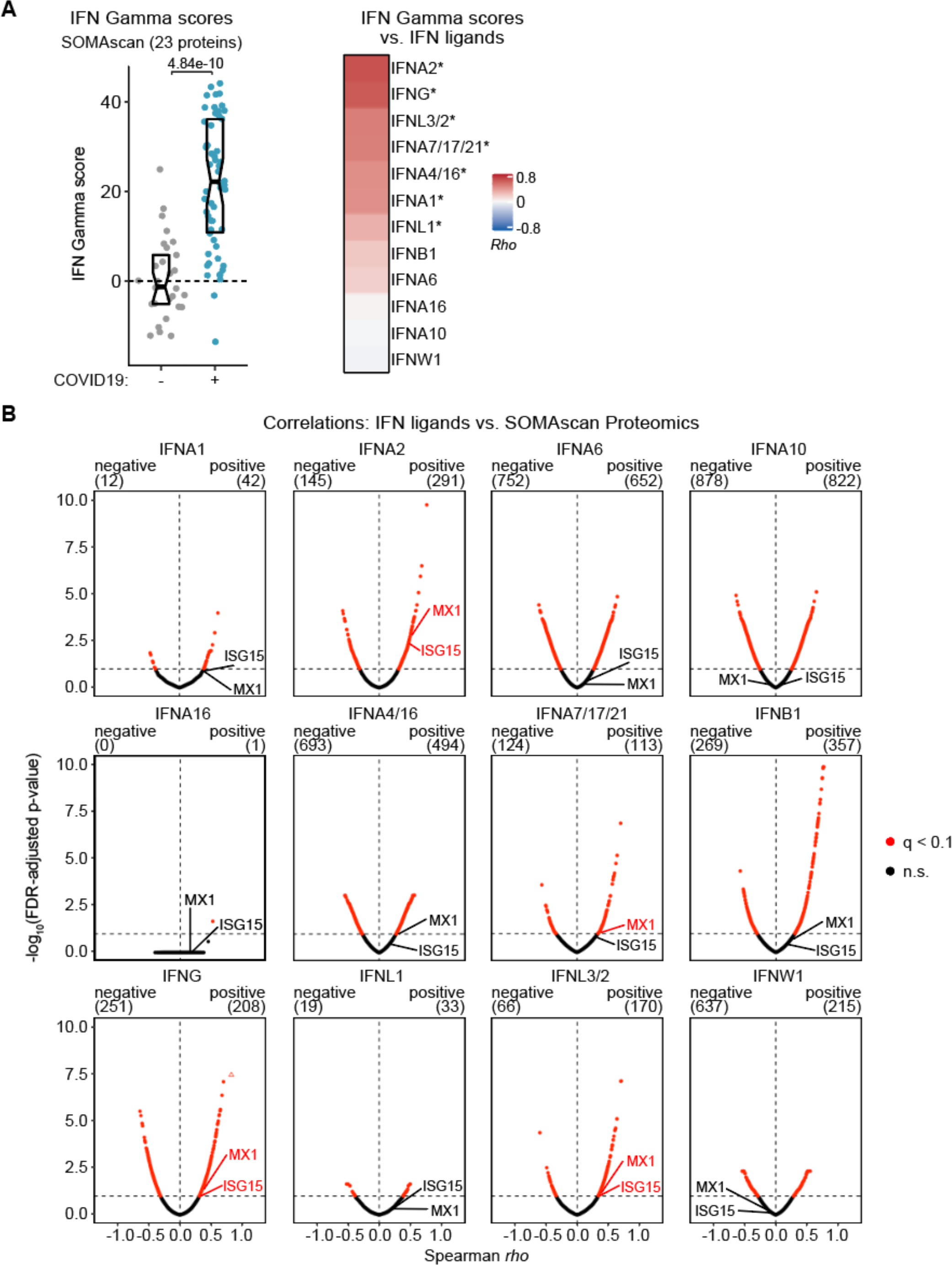
Plasma protein-based IFN Gamma scores and correlation analysis for IFN ligands vs. SOMAscan^®^ proteomics. (**A**) Sina plot of protein-based IFN Gamma scores, separated by COVID19 status and ranked heatmap representing correlations between protein-based IFN Gamma scores and plasma levels of each IFN ligand. IFN gamma scores were calculated for each research participant by summing Z-scores for 23 differentially abundant proteins from the IFN Gamma Response Hallmark gene set from MSigDB. Z-scores were calculated from the adjusted concentration values for each gene in each sample, based on the mean and standard deviation of COVID19-negative samples. Data are presented as a modified sina plot with box indicating median and interquartile range. Heatmap values displayed are Spearman correlation coefficients (Rho); asterisks indicate significant correlations (10% FDR). (**B**) Volcano plots for Spearman correlation analysis of IFN ligands vs. Somascan protein abundance values. Horizontal dashed line indicates an FDR threshold of 10% (q < 0.1); red points and numbers above plots indicate significant proteins at this threshold.

**Figure 2 – supplement 2.**
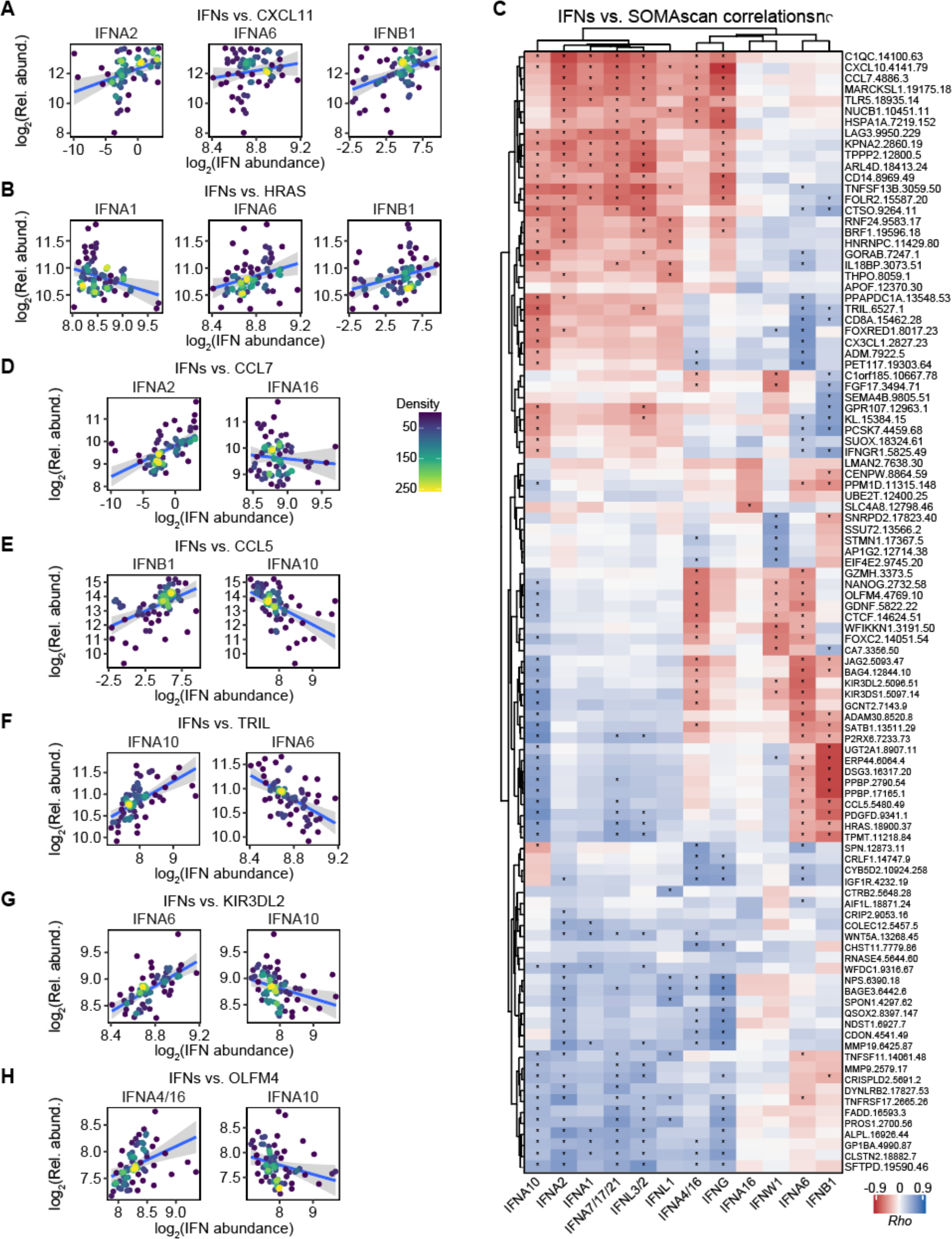
Relationships between IFN ligands and SOMAscan^®^ plasma proteomics. (**A-B** and **D-H**) Scatter plots comparing relationships between plasma proteins and the indicated IFNs in COVID19-positive patients. Points are colored by density; blue lines represent linear model fit with 95% confidence intervals in grey. (C) Heatmap representing correlations between plasma levels of proteins measured by SOMAscan^®^ and each IFN ligand. Values displayed are Spearman correlation scores (Rho) for proteins ranked in top 5 positive or top 5 negative correlations for at least one IFN; asterisks indicate significant correlations (10% FDR); columns and rows are grouped by hierarchical clustering.

**Figure 3 – supplement 1.**
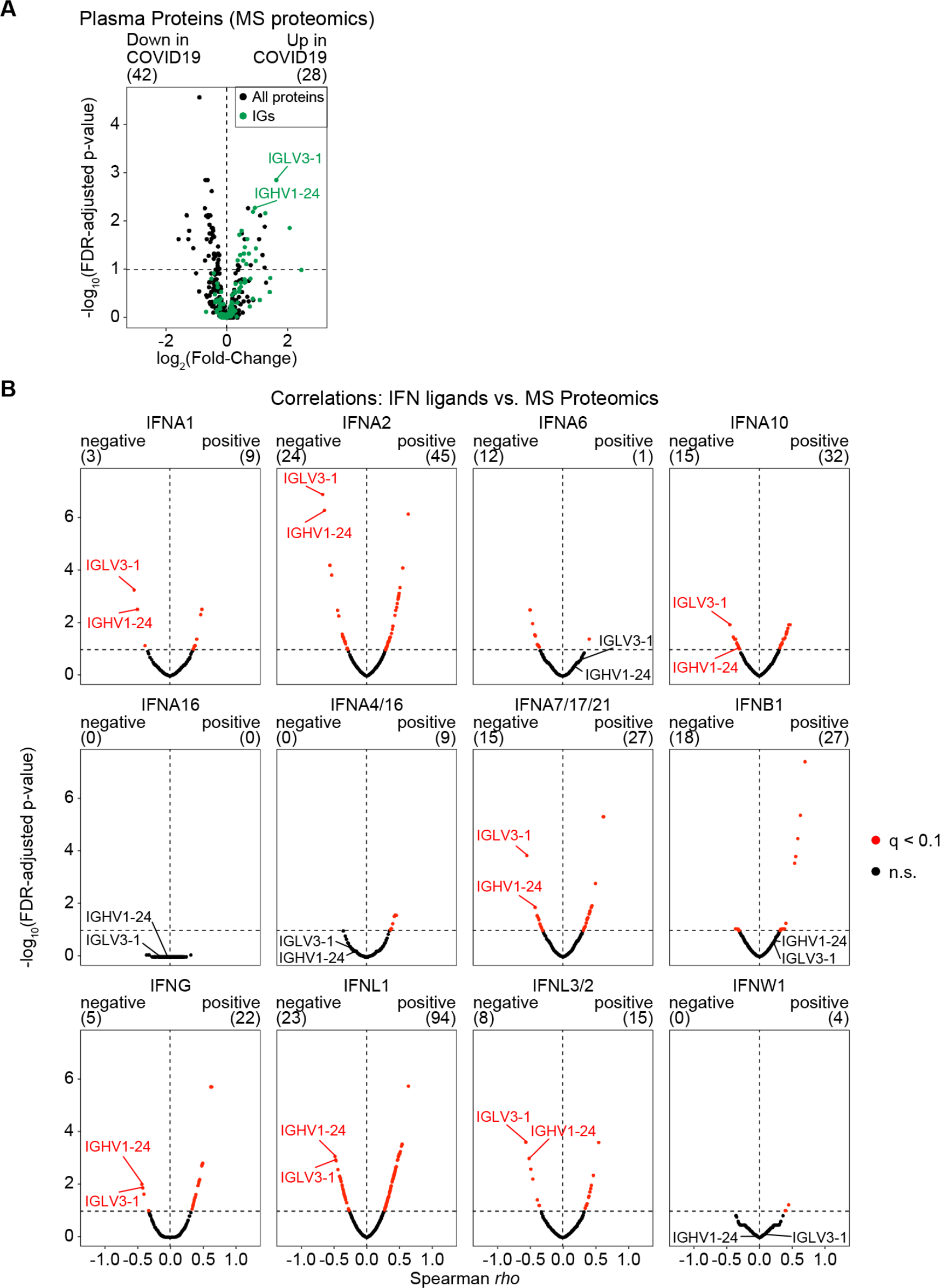
Differential abundance and correlation analysis for MS plasma proteomics. (**A**) Volcano plot for linear regression analysis of MS proteomics plasma protein abundance data for COVID19-positive vs. -negative samples, adjusted for age and sex. Horizontal dashed line indicates an FDR threshold of 10% (q < 0.1); numbers above plot indicate significant genes at this threshold. Immunoglobulin subunits (IGs) are highlighted in green. (**B**) Volcano plots for Spearman correlation analysis of IFN ligands vs. MS proteomics protein relative abundance values. Horizontal dashed line indicates an FDR threshold of 10% (q < 0.1); red points and numbers above plots indicate significant proteins at this threshold.

**Figure 3 – supplement 2.**
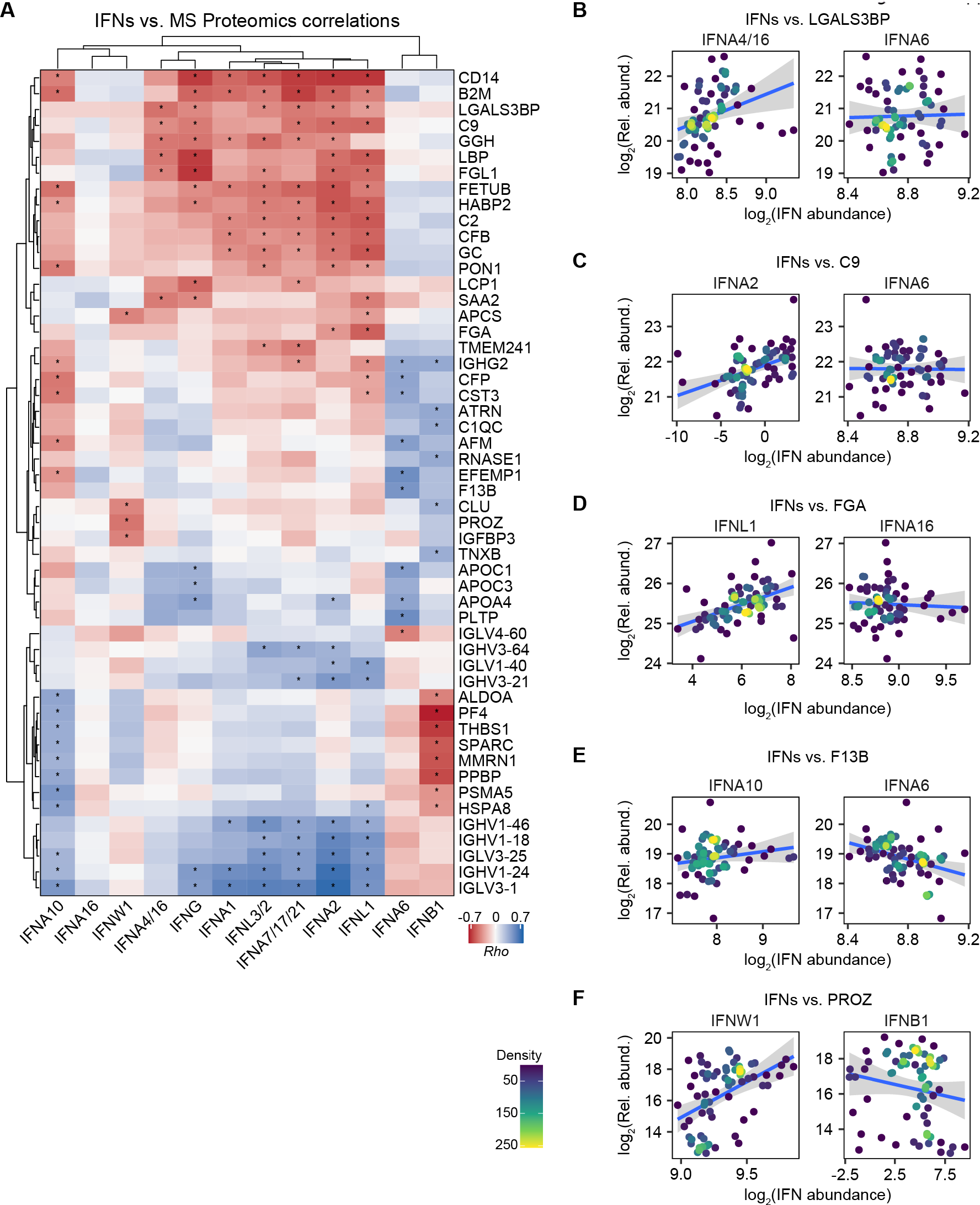
Relationships between IFN ligands and MS plasma proteomics. (**A**) Heatmap representing correlations between IFN ligands and plasma protein levels, as measured by MS proteomics. Values displayed are Spearman correlation scores (Rho) for proteins ranked in top 5 positive or top 5 negative correlations for at least one IFN; asterisks indicate significant correlations (10% FDR); columns and rows are grouped by hierarchical clustering. (**B-F**) Scatter plots comparing relationships between plasma proteins, as measured by MS proteomics, and the indicated IFNs in COVID19-positive patients. Points are colored by density; blue lines represent linear model fit with 95% confidence intervals in grey.

**Figure 4 – supplement 1.**
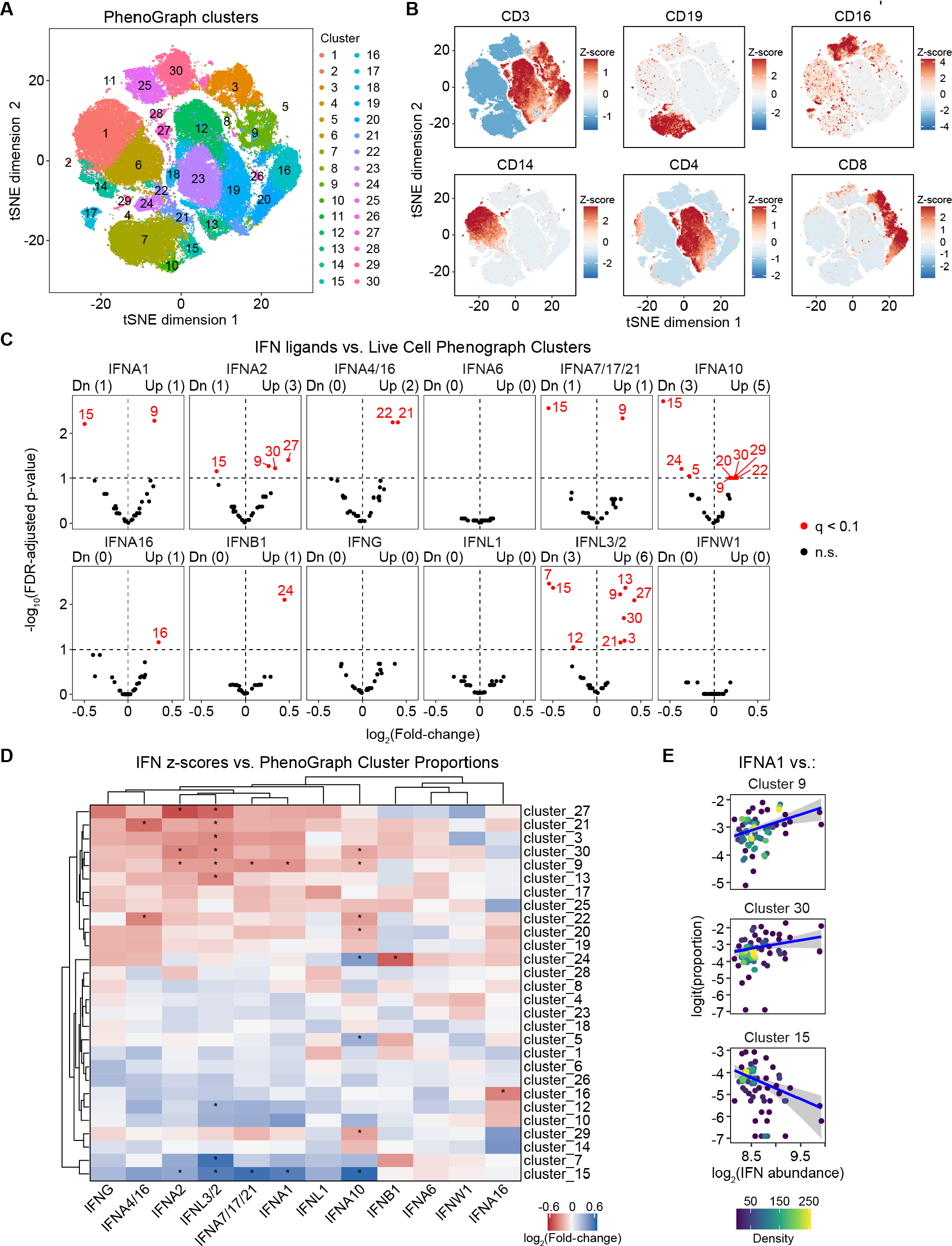
PhenoGraph clustering and beta regression analysis of clustered mass cytometry data against IFN ligands. (**A**) t-SNE plots of 69,000 cells analyzed by mass cytometry from 69 COVID19-positive patients (1,000 cells each). Numbers and coloring of cells indicate PhenoGraph cluster assignments. (**B**) t-SNE plots with cells colored by Z-scores for markers of T cells (CD3, CD4, CD8), B cells (CD19), NK cells (CD16), and Monocytes (CD14). (**C**) Volcano plots for Beta regression analysis of IFN ligands vs. cluster proportions among live cells, adjusted for age and sex. X-axes display log2-transformed fold-change in cluster proportion among live cells per standard deviation of IFN abundance; horizontal dashed line indicates an FDR threshold of 10% (q < 0.1); red points and numbers above plots indicate significant clusters at this threshold. (**D**) Heatmap representing relationships between IFN ligands and cluster proportions among live cells, as determined by beta regression analysis. Values displayed are fold-change in cluster proportion among live cells per standard deviation of IFN abundance; asterisks indicate significant associations (10% FDR); columns and rows are grouped by hierarchical clustering. (**E**) Scatter plots comparing relationships between cluster proportions among live cells, and the indicated IFNs in COVID19-positive patients. Points are colored by density; blue lines represent beta regression model fit with 95% confidence intervals in grey.

**Figure 4 – supplement 2.**
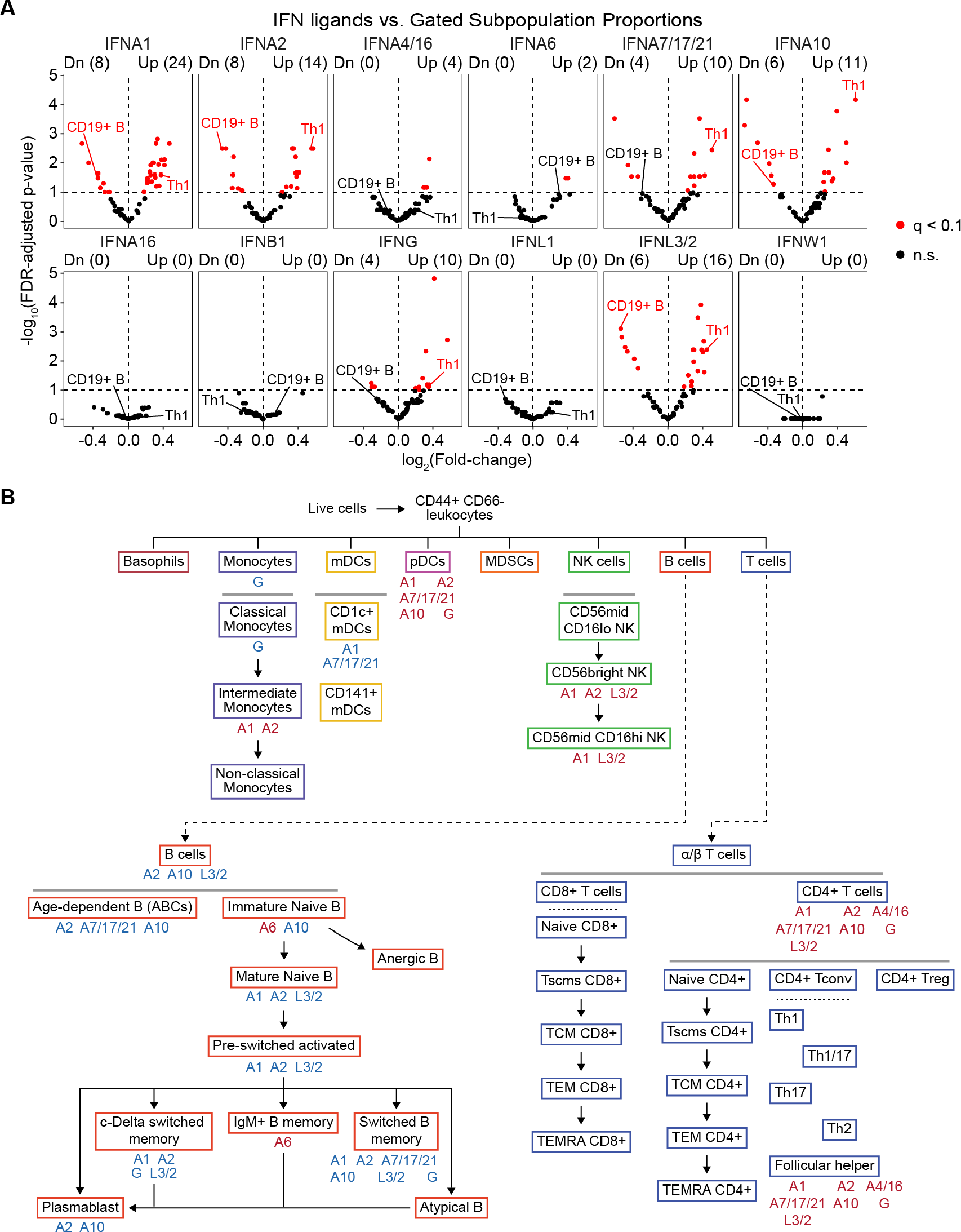
Beta regression analysis of gated mass cytometry data against IFN ligands. (**A**) Volcano plots for Beta regression analysis of IFN ligands vs. gated subpopulation proportions among live cells, adjusted for age and sex. X-axes display log2-transformed fold-change in cluster proportion among live cells per standard deviation of IFN abundance; horizontal dashed line indicates an FDR threshold of 10% (q < 0.1); red points and numbers above plots indicate significant subpopulations at this threshold. (**B**) Cell lineage map indicating the relationships between gated cell subpopulations included in beta regression analysis. Boxed labels represent subpopulations for which relative cell frequencies were obtained. Grey horizontal lines denote subpopulations of cells derived by gating; black lines and arrows indicate subpopulations that are also related by cell differentiation; labels outside boxes indicate significant positive (red) and negative (blue) relationships with IFN ligands.

**Figure 5 – supplement 1.**
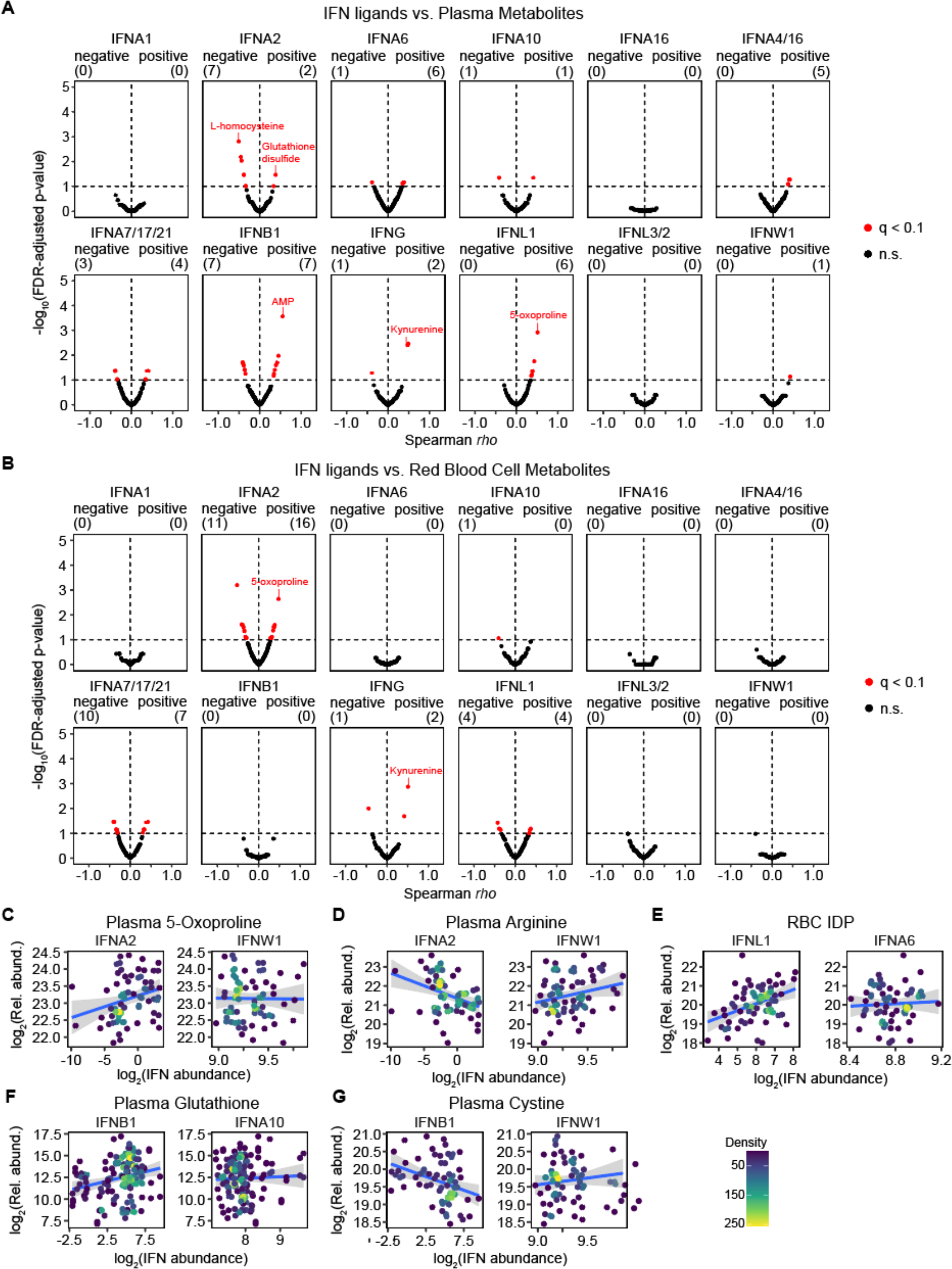
Correlation analysis of plasma and RBC metabolites vs. IFN ligands. (**A-B**) Volcano plots for Spearman correlation analysis of IFN ligands vs. plasma (A) or RBC (B) metabolite relative abundance values. Horizontal dashed line indicates an FDR threshold of 10% (q < 0.1); red points and numbers above plots indicate significant metabolites at this threshold. (**C-G**) Scatter plots comparing relationships between select metabolites and the indicated IFNs in COVID19-positive patients. Points are colored by density; blue lines represent linear model fit with 95% confidence intervals in grey.

